# Cross omics risk scores of inflammation markers are associated with all-cause mortality: The Canadian Longitudinal Study on Aging

**DOI:** 10.1101/2024.09.24.24313672

**Authors:** Anat Yaskolka Meir, Huan Yun, Jie Hu, Jun Li, Jiaxuan Liu, Alaina Bever, Andrew Ratanatharathorn, Mingyang Song, A. Heather Eliassen, Lori Chibnik, Karestan Koenen, Guillaume Pare, Meir J Stampfer, Liming Liang

## Abstract

Inflammation is a critical component of chronic diseases, aging progression, and lifespan. Omics signatures may characterize inflammation status beyond blood biomarkers. We leveraged genetics (Polygenic-Risk-Score; PRS), metabolomics (Metabolomic-Risk-Score; MRS), and epigenetics (Epigenetic-Risk-Score; ERS) to build multi-omics-multi-marker risk scores for inflammation status represented by the level of circulating C-reactive protein (CRP), interleukin 6 (IL6), and tumor necrosis factor alpha (TNFa). We found that multi-omics risk-scores generally outperformed single-omics risk scores in prediction of all-cause mortality in the Canadian Longitudinal Study on Aging. Compared with circulating inflammation biomarkers, some multi-omics risk scores had a higher HR for all cause-mortality when including both score and circulating IL6 in the same model (1-SD IL6 MRS-ERS: HR=1.77 [1.15-2.72] vs. 1-SD circulating IL6 HR=1.11 [0.75,1.66]; 1-SD IL6 PRS-MRS: HR=1.32 [1.21,1.45] vs. 1-SD circulating IL6 HR=1.31 [1.12, 1.53]; 1-SD PRS-MRS-ERS: HR=1.62 [1.04, 2.53] vs. 1-SD circulating IL6: HR=1.16 [0.77, 1.74]). In the Nurses’ Health Study (NHS), NHS II, and Health Professional Follow-up Study with available omics, 1-SD of IL6 PRS and 1-SD IL6 PRS-MRS had HR=1.13 [1.00,1.27] and HR=1.13 [1.01,1.27], among individuals >65years without mutual adjustment of the score and circulating IL6. Our study demonstrated that some multi-omics scores for inflammation markers may characterize important inflammation burden for an individual beyond those represented by blood biomarkers and improve our prediction capability for aging process and lifespan.

## Introduction

Strong evidence supports the role of low-grade systemic inflammation in complex diseases, including cardiovascular disease and type 2 diabetes ^1–3^, as well as age-related diseases such as Alzheimer’s disease (AD) ^4^. Aging and age-related diseases are associated with a decline in immune function, known as immunosenescence, and an increase in inflammation, sometimes referred to as “inflammaging” ^5^. Immunosenescence is associated with an imbalance of pro and anti-inflammatory cytokines, and a chronic low-grade inflammatory state has been linked to poor function and mobility in older adults ^5^. Among traditionally measured inflammation biomarkers, blood C-reactive protein (CRP), interleukin 6 (IL6), and tumor necrosis factor alpha (TNFa) levels are independent predictors for all-cause mortality in several prospective studies ^6^ ^7,8^, especially in those above 65 years ^9^ ^8^. However, a snapshot of these biomarkers from the blood may not reflect the full inflammation status of an individual.

Integrating different omics reflecting different functional layers (“multi-omics”) may better characterize a more comprehensive spectrum of inflammation and the burden on chronic inflammation from immediate and acute status (e.g., metabolomics) to lifetime impact (e.g., genetics). Using multi omics jointly may enhance our ability to evaluate and explore inflammatory-related pathophysiology.

In this study, leveraging data from the Canadian Longitudinal Study on Aging (CLSA) and published genome-wide summary statistics, we established multi-omics scores for inflammation markers CRP, IL6, and TNFa based on genetics (via Polygenic Risk Score or PRS), metabolomics (via Metabolomic Risk Score or MRS), and epigenetics (via Epigenetic Risk Score or ERS, using DNA methylation). We hypothesized that, independent of observed level of blood biomarkers, individuals with an omics-based inflammation signatures of CRP, IL6 or TNFa levels would carry a greater burden of chronic inflammation and therefore have a higher hazard of mortality. The risk scores and the association with all-cause mortality were further tested for validation in the Nurses’ Health Study (NHS), NHS II, and the Health Professional Follow-up Study (HPFS).

## Methods

### Study population

We used the comprehensive CLSA cohort (N=30,097 participants; 50.9% women, mean (SD) age=62.96 (10.25)), which randomly selected from within 25–50 km of 11 data collection sites in 7 Canadian provinces and were interviewed in person, took part in in-depth physical assessments at the collection sites, and provided blood and urine samples (2011-2015)^10^. The first (2015-2018) and the second (2018-2021) follow-ups on the comprehensive cohort includes reports on all-cause mortality. The CLSA data were used to train the metabolomic and epigenetic prediction models and for the discovery association study of the omics-based signatures with all-cause mortality. The NHS, NHS II, and HPFS were used to validate the risk scores and the association with mortality. The description of the NHS, NHS II, and HPFS is presented in **Methods S1**.

### Assessment of omics

Details on measuring and processing the omics data in the CLSA are presented below. The methods for omics in the validation cohorts are described in **Methods S2**.

#### Genotyping and imputation

The DNA extraction, genotyping, and quality filtering protocol described previously^11^ ^12^. Briefly, genotyping was undertaken in 5 batches of roughly 5,000 samples, each using Axiom™ Analysis Suite 2.0, similar to the UK Biobank genotyping QC documentation^13^. The average call rate for passing samples was ≥95.0 and the Hardy-Weinberg equilibrium (HWE) p-value was > 10^−6^. Imputation quality using the TOPMed reference panel was assessed using the marker-wise information measure (Rsq) and compared to the imputation using the Haplotype Reference ^14^ . SNPs with minor allele frequency (MAF) < 0.05 were removed. Out of 794,409 genetic markers, 573,386 were selected after sample-based quality control. The above procedure to remove duplicates resulted in 26,622 uniquely genotyped CLSA participants.

#### Metabolomics

Blood metabolomics were evaluated for 9,992 samples at baseline at Metabolon, Inc. Quality control and normalization methods are detailed in the CLSA Metabolomic Profiling Data Support Document^15^. For the current study, the following metabolite groups were included: Amino acids, Xenobiotics, Peptides, Partially Characterized Molecules, Nucleotides, Lipids, Energy, Cofactors and Vitamins, and Carbohydrates (a total of 1030 metabolites). Metabolites were further standardized to a mean of 0 and SD of 1 before inputting in the models.

#### DNA methylation profiling

The site-specific DNA methylation was measured using the Infinium MethylationEPIC BeadChip platform (Illumina, CA, USA) on DNA extracted from peripheral blood mononuclear cells (PBMCs) as previously described ^16^ ^17^. For this study, we performed sample-level quality control (QC) using functions from the R package “minfi” ^18^. We evaluated the quality of 1478 samples at baseline by: i. Computing the median for both Meth and Unmeth signals for each array and displaying in a scatter plot to identify outlier samples with low intensity. The cutoff of the median log2 intensity value was < 10.5; ii. Evaluating outlier samples and confirming male and female blood samples clustered separately according to Multi-Dimensional Scaling (MDS) plots. This was repeated twice: clustering by the predicted sex and clustering by the reported sex. Two samples were flagged: one sample had a predicted sex different from the reported sex, and one sample clustered within the opposite sex that was predicted and reported; iii. Calculating sample-wise missing rates. We set a detection p-value of 0.01 and > 5% missing rate as a threshold to be removed. iv. Additional quality control was performed using the QC report generated by “qcReport” function of the minfi R package. For the Probe-level QC, we used the “preprocessQuantile” function to normalize the data and “getBeta” function to get the final beta values for 1476 samples.

### Assessment of inflammation markers

Circulating inflammatory biomarker measurements were detailed before ^19^. Briefly, CRP for 27,011 participants was measured in serum using the Cobas 8000 modular analyzer (Roche Diagnostics). TNFa and IL6 (available for N=9,522 and N=9,698 participants) were measured in serum using the Quantikine high sensitivity ELISA (R & D Systems). Blood draws for inflammation markers were performed at the same time as the omics data at baseline. The current analysis included participants with overlapping blood inflammation markers and omics data.

Measurement of blood CRP and IL6 in the validation cohorts NHS, NHS II, and HPFS is described in **Methods S3**. For the NHS/HPFS, we used inflammation markers measured closest to the timing of blood draws for omics. Of note, blood TNFa was not measured in these studies.

### Main covariates

Background characteristics used for the all-cause mortality association study were age, sex, smoking status (yes/no/former), BMI, alcohol intake (frequency of alcohol consumption past 12 months), and race/ethnicity, which were collected at baseline. Multi-morbidity was defined as an ordinal measurement, ranging from 0 to 8, and calculated by summing the number of the following diseases in the CLSA (assessed by self-reports at baseline): Dementia or Alzheimer’s disease, Parkinson’s disease, multiple sclerosis, diabetes mellites, renal failure, musculoskeletal system, and connective tissue disease (osteoporosis, rheumatoid arthritis, arthritis), cardiovascular risk factors and disease (hypertension, heart disease including congestive heart failure, angina, myocardial infarction or heart attack, coronary artery bypass graft surgery (CABG) surgery or percutaneous coronary intervention, congestive heart failure, stroke), and cancer (except non-melanoma skin cancer). The diseases included in the multimorbidity were based on previous publications^20,21^ .

### Prediction models

#### Single-omics (1-way) risk scores

PRS for inflammation was detailed previously ^22^. Briefly, GWAS for CRP, IL6, and TNFa, was curated from summary data from several publicly available sources. We conducted a meta-analysis for the same circulating protein biomarkers using METAL^23^ with the inverse-variance-weighted method. Using the genome-wide SNPs from the meta-analysis summary statistics, PRS was generated by applying SBayesian linear mixed models with LD Sparse matrix (ukbEURu_imp_v3_HM3_n50k.chisq10.ldm.sparse) as demonstrated before ^24^. For MRS and ERS models, we first applied log10 transformation on the blood inflammation markers to reduce skewness and control the effect from outliers. We generated MRS and ERS for each inflammation marker separately using Elastic net regression (“glmnet” R package) with all metabolites and DNA methylation sites as independent variables. To reduce over-fitting, a 10-fold cross-validation (CV) was used to obtain the predicted value for each CLSA individual. Pearson R and root mean square error (RMSE) were calculated based on the observed and the predicted values. The coefficients used on the validation sets were generated from the last fold.

#### Multi-omics (2- and 3-way) risk scores

we established the risk scores based on each omics pair combination (PRS-ERS, PRS-MRS, MRS-ERS) or all three (PRS-MRS-ERS) using a hierarchical approach. In this approach, we first used the omics with the largest number of samples and regress the corresponding omics’ score out of the blood biomarker by linear regression. Next, we input the residual from this linear regression as the outcome in an elastic net regression with the second-largest omics as the predictors. For 2-way risk scores, we sum the predictors from the second step and the prediction variable by risk score*beta from the first linear regression. For the 3-way risk scores, we build additional regression predicting the inflammation marker by the previous two omics scores and use the residuals as an outcome for another 10-fold CV elastic net regression with the third set of omics as predictors. The 3-way risk score is the sum of beta from the second linear regression*PRS + beta from the second linear regression*precited by MRS + risk score output from the final elastic net (ERS). The conceptual framework for establishing 1-, 2- and 3-way risk scores is detailed in **Figure 1**.

**Figure 1:**
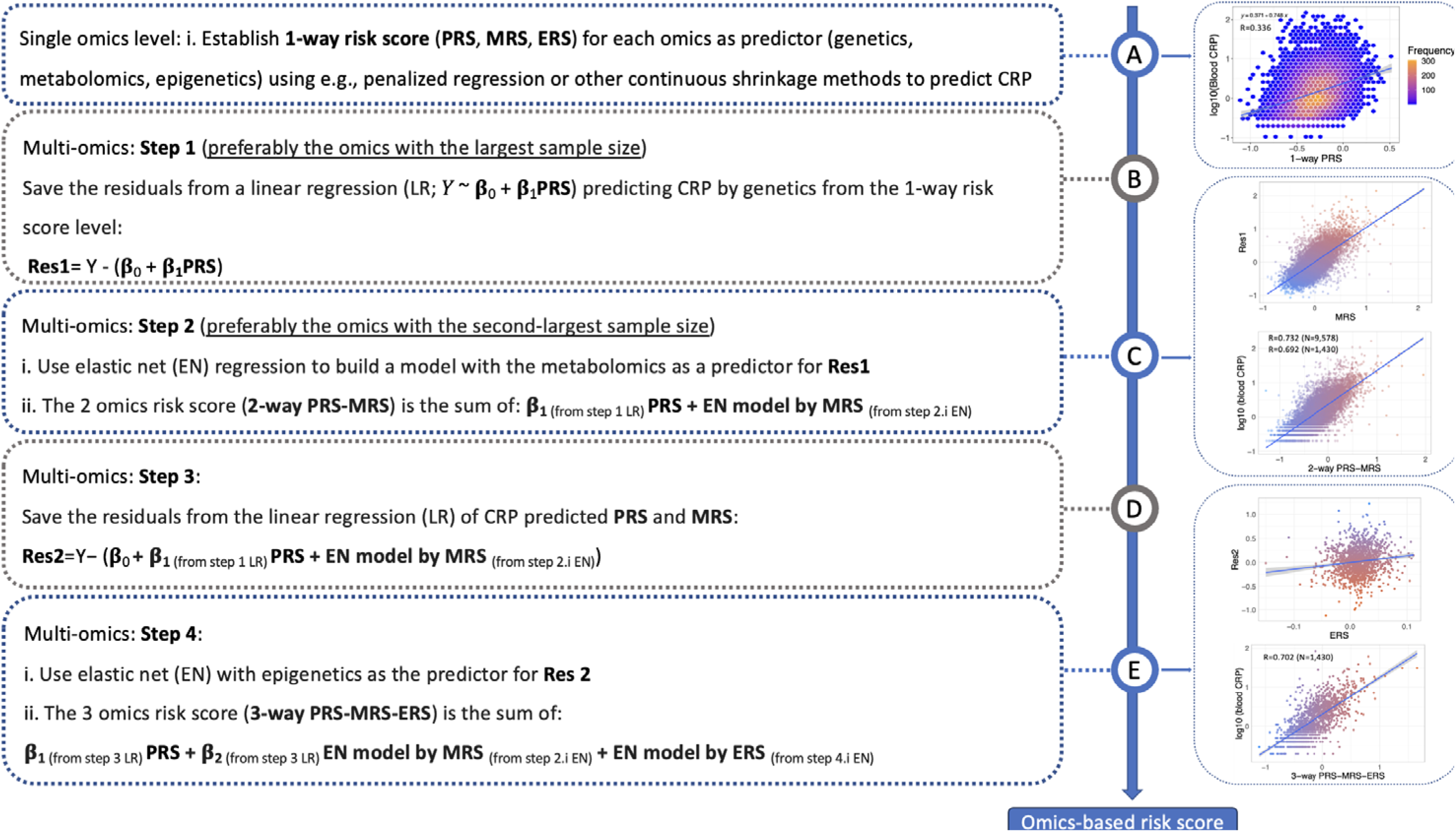
Analyses workflow and application for 1-, 2-, and 3-way risk scores and hierarchical approach for multi-omics integration. Steps for establishing omics risk scores using genetics, metabolomics, and DNA methylation demonstrated for blood CRP.

### Statistical analysis

Blood CRP, IL6, and TNFa were Log10 transformed before being used in all models and correlation tests. Pearson correlation was used to examine the correlation between the observed log-transformed inflammation markers and the omics risk scores. Spearman correlation was used to examine correlations between inflammation markers and age. We applied previously published DNA methylation scores for CRP and IL6 onto the CLSA data: i. The CRP score by Ligthart 2016 ^25^, suggested using 218 CpGs to predict CRP (207 available in the CLSA data); ii. The score by Barker 2018 ^26^ included 7 CpGs (6 available) to predict CRP; iii. The score by Stevenson 2021^27^ included 35 CpGs (all available) to predict IL6. The association with all-cause mortality was examined using Cox regression (“survival” R package), with the follow-up time from baseline to either the death or the end of the follow-up period. The models for the survival discovery analysis were adjusted for age, sex, smoking, alcohol intake, body mass index (BMI), race/ethnicity, and multi-morbidity. We reported the Hazard Ratios (HR), 95% confidence intervals (CI), and concordance resulting from the Cox models. We calculated the survival models’ dynamic Area Under the Curve (AUC) using the “dynpred “R package. Sensitivity analyses were performed using Logistic regression, using the same covariates as in the Cox models. The risk scores and blood inflammation markers were standardized before input to the models, with a mean of 0 and SD of 1. Thus, the effect size of the Cox and Logistic regression models represents a 1-SD change of each of these predictors. Likelihood ratio test (LRT) was performed using the “lrtest” function in the “lmtest” R package. All statistical analyses were performed using R (version 4.2.3; R Foundation for Statistical Computing).

## Results

### Description of cohorts used for this analysis

The background characteristics of the CLSA cohort are presented in **Table 1**. No significant differences in terms of age and sex were observed between participants with metabolomics and epigenetics data and those without omics data (all p>0.05). However, participants without genetic data (N=3,475) were slightly older (63.5 years vs. 62.9 years) and with a higher proportion of women (58.7% vs. 49.9%) compared with participants with genetic data (N=26,622). The three inflammation markers CRP, IL6, and TNFa were directly correlated with age (CRP: R= 0.105; IL6: R= 0.279; TNFa: R= 0.289; p<0.001 for all; **Figure S1**). The characteristics of the validation sets (NHS II: PRS, MRS, ERS; NHS and HPFS: PRS and MRS) are presented in **Table S1**.

**Table 1:** Background characteristics of the CLSA across the availability of omics data

| Characteristics | Individuals with genetic data<br>N=26,622 | Individuals with metabolomic data<br>N=9,992 | Individuals with epigenetic data<br>N=1,478 |
| --- | --- | --- | --- |
| Age, years | 62.88 (10.20) | 63.00 (10.20) | 63.12 (10.29) |
| Men, % | 50.12% | 49.10% | 49.46% |
| Body mass index, kg/m<sup>2</sup> | 28.07 (8.02) | 28.07 (5.44) | 28.43 (5.69) |
| Current smokers, % | 8.93% | 9.45% | 9.95% |
| CRP, mg/L<sup>1</sup> | 2.56 (5.07) | 2.54 (4.69) | 2.68 (4.42) |
| IL6, pg/mL<sup>2</sup> | 2.38 (1.63) | 2.38 (1.63) | 2.62 (1.74) |
| TNFa, pg/mL<sup>3</sup> | 1.10 (0.50) | 1.11 (0.50) | 0.49 (7.16) |

\*Sample size before QC steps. Data is presented as means (SD) for continuous measurements and numbers/percentages for categorical. <sup>1</sup> N with available data: genetic sample: 26,462; metabolomic sample: 9,958; epigenetic sample: 1,470. <sup>2</sup> N with available data: genetic sample: 9,537; metabolomic sample: 9,728; epigenetic sample: 1,420. <sup>3</sup> N with available data: genetic sample: 9,360; metabolomic sample: 9,954; epigenetic sample: 1,428.

### One-way omics risk scores for inflammation markers

#### Inflammation PRS

We applied our pre-determined inflammation PRS^22^ to the CLSA, NHS, NHS II, and HPFS. In the CLSA, CRP PRS had the strongest correlation with blood CRP levels (N=26,462, R=0.336, p<0.001). PRS for IL6 and TNFa correlations with the corresponding blood levels were significant but weaker (N=9,537, R=0.049, p<0.001 and N=9,360, R=0.032, p=0.001; for IL6 and TNFa, respectively; **Tables 2-4**, **Figure S2a-c**). Similar correlations between blood CRP and CRP PRS were observed in the NHS, NHS II, and HPFS (**Table 5**). Blood IL6 was strongly correlated with IL6 PRS in all validation cohorts (**Table 5**).

**Table 2:** Multi-omics inflammation signature for CRP and association with all-cause mortality in CLSA

| N omics | Approach | The number and type of predictors included in the final model | Sample size to calculate model performance | RMSE* | R (Pearson) | Analysis model | 1-SD of blood CRP | 1-SD of the signature | Sample size and number of events** |
| --- | --- | --- | --- | --- | --- | --- | --- | --- | --- |
| 3-way: PRS-MRS-ERS | Hierarchical | CRP predicted by 1 PRS, CRP predicted by 12 CpGs, CRP predicted by 308 metabolites | 1,430 | 0.442 | 0.703 | Age, sex, smoking, multimorbidity, BMI, race/ethnicity, and alcohol-adjusted model (MV) | 1.35<br>(0.95,1.90) | **1.64**<br>**(1.13,2.37)** | N=946; number of events=43 |
|  |  |  |  |  |  | MV + and mutual adjustment | 1.05<br>(0.68,1.61) | 1.59 (0.99,2.54) |  |
| 2-way: PRS-MRS | Hierarchical | 1 PRS + predicted value by 308 metabolites | 9,758<br>[1,430] | 0.474<br>[0.495] | 0.732<br>[0.692] | Age, sex, smoking, multimorbidity, BMI, race/ethnicity, and alcohol-adjusted model (MV) | **1.29**<br>**(1.14, 1.46)** | **1.32**<br>**(1.15,1.51)** | N=6,521; number of events=275 |
|  |  |  |  |  |  | MV + and mutual adjustment | 1.16<br>(0.98;1.38) | 1.19<br>(0.99, 1.42) |  |
| 2-way: PRS-ERS | Hierarchical | 1 PRS + predicted value by 428 CpGs | 1,435 | 0.514 | 0.577 | Age, sex, smoking, multimorbidity, BMI, race/ethnicity, and alcohol-adjusted model (MV) | **1.40**<br>**(1.01,1.93)** | 1.41 (0.98,2.03) | N=950; number of events=45 |
|  |  |  |  |  |  | MV + and mutual adjustment | 1.28<br>(0.89,1.84) | 1.28 (0.86,1.88) |  |
| 2-way: MRS-ERS | Hierarchical | predicted value by 47 CpGs + predicted value by 319 metabolites | 1,463 | 0.323 | 0.664 | Age, sex, smoking, multimorbidity, BMI, race/ethnicity, and alcohol-adjusted model (MV) | 1.34<br>(0.95, 1.87) | **1.76**<br>**(1.22, 2.53)** | N=966; number of events=45 |
|  |  |  |  |  |  | MV + and mutual adjustment | 0.99<br>(0.66, 1.49) | **1.77**<br>**(1.14, 2.77)** |  |
| 1-way: PRS | PRS-CS | 1 PRS | 26,462 | 0.602 | 0.336 | Age, sex, smoking, multimorbidity, BMI, race/ethnicity, and alcohol-adjusted model (MV) | **1.25**<br>**(1.16, 1.34)** | 1.02 (0.95,1.10) | N=17,612; number of events=759 |
|  |  |  |  |  |  | MV + and mutual adjustment | **1.28**<br>**(1.18,1.38)** | 0.94 (0.87,1.01) |  |
| 1-way: MRS | 10-fold CV elastic net | 319 metabolites | 9,956 | 0.315 | 0.688 | Age, sex, smoking, multimorbidity, BMI, race/ethnicity, and alcohol-adjusted model (MV) | **1.30**<br>**(1.14, 1.47)** | **1.43 (1.26,1.62)** | N=6,660; number of events=284 |
|  |  |  |  |  |  | MV + and mutual adjustment | 1.10<br>(0.94,1.28) | **1.35**<br>**(1.15, 1.59)** |  |
| 1-way: ERS | 10-fold CV elastic net | 150 CpGs | 1,468 | 0.371 | 0.534 | Age, sex, smoking, multimorbidity, BMI, race/ethnicity, and alcohol-adjusted model (MV) | 1.38<br>(0.99,1.92) | **1.64 (1.22,2.21)** | N=970; number of events=47 |
|  |  |  |  |  |  | MV + and mutual adjustment | 1.21<br>(0.85,1.71) | **1.56**<br>**(1.13, 2.13)** |  |

\*RMSE for 2- and 3-way models was calculated as follows: $RMSE = \sqrt{\sum (P_i - O_i)^2 / n}$ . \*\*According to MV + mutual adjustment. Bold values denote $p < 0.05$

**Table 3:** Multi-omics inflammation signature for IL6 and association with all-cause mortality in CLSA

| N omics | Approach | The number and type of predictors included in the final model | Sample size to calculate model performance | RMSE* | R (Pearson) | Analysis model | 1-SD of blood CRP | 1-SD of the signature | Sample size and number of events** |
| --- | --- | --- | --- | --- | --- | --- | --- | --- | --- |
| 3-way: PRS-MRS-ERS | Hierarchical | IL6 predicted by 1 PRS, IL6 predicted by 37 CpGs, IL6 predicted by 290 metabolites | 1384 | 0.368 | 0.620 | Age, sex, smoking, multimorbidity, BMI, race/ethnicity, and alcohol-adjusted model (MV) | 1.43<br>(0.99, 2.06) | **1.75**<br>**(1.18, 2.59)** | N=920; number of events=40 |
|  |  |  |  |  |  | MV + and mutual adjustment | 1.16<br>(0.77, 1.74) | **1.62**<br>**(1.04, 2.53)** |  |
| 2-way: PRS-MRS | Hierarchical | 1 PRS + predicted value by 290 metabolites | 9533<br>[1384] | 0.3688<br>[0.393] | 0.647<br>[0.609] | Age, sex, smoking, multimorbidity, BMI, race/ethnicity, and alcohol-adjusted model (MV)) | **1.50**<br>**(1.30, 1.74)** | **1.38**<br>**(1.28, 1.48)** | N=6403; number of events=260 |
|  |  |  |  |  |  | MV + and mutual adjustment | **1.31**<br>**(1.12, 1.53)** | **1.32**<br>**(1.21, 1.45)** |  |
| 2-way: PRS-ERS | Hierarchical | 1 PRS + 549 CpGs | 1385 | 0.367 | 0.565 | Age, sex, smoking, multimorbidity, BMI, race/ethnicity, and alcohol-adjusted model (MV) | **1.47**<br>**(1.03, 2.10)** | 1.34<br>(0.90, 2.00) | N=921; number of events=40 |
|  |  |  |  |  |  | MV + and mutual adjustment | 1.34<br>(0.89, 2.02) | 1.17<br>(0.76, 1.81) |  |
| 2-way: MRS-ERS | Hierarchical | predicted value by 130 CpGs and predicted value by 232 metabolites | 1417 | 0.211 | 0.619 | Age, sex, smoking, multimorbidity, BMI, race/ethnicity, and alcohol-adjusted model (MV) | **1.43**<br>**(1.00, 2.03)** | **1.87**<br>**(1.29, 2.72)** | N=940; number of events=42 |
|  |  |  |  |  |  | MV + and mutual adjustment | 1.11<br>(0.75, 1.66) | **1.77**<br>**(1.15, 2.72)** |  |
| 1-way: PRS | PRS-CS | 1 PRS | 9,360 | 0.664 | 0.049 | Age, sex, smoking, multimorbidity, BMI, race/ethnicity, and alcohol-adjusted model (MV) | **1.49**<br>**(1.28, 1.73)** | 0.94<br>(0.83, 1.06) | N=6406; number of events=260 |
|  |  |  |  |  |  | MV + and mutual adjustment | **1.49**<br>**(1.29, 1.73)** | 0.93<br>(0.83, 1.05) |  |
| 1-way: MRS | 10-fold CV elastic net | 232 metabolites | 9,727 | 0.206 | 0.656 | Age, sex, smoking, multimorbidity, BMI, race/ethnicity, and alcohol-adjusted model (MV)) | **1.51**<br>**(1.30, 1.74)** | **1.63**<br>**(1.45, 1.83)** | N=6539; number of events=269 |
|  |  |  |  |  |  | MV + and mutual adjustment | **1.24**<br>**(1.05, 1.46)** | **1.50**<br>**(1.30, 1.74)** |  |
| 1-way: ERS | 10-fold CV elastic net | 469 CpGs | 1,418 | 0.220 | 0.575 | Age, sex, smoking, multimorbidity, BMI, race/ethnicity, and alcohol-adjusted model (MV) | **1.43**<br>**(1.00, 2.03)** | **1.59**<br>**(1.09, 2.32)** | N=941; number of events=42 |
|  |  |  |  |  |  | MV + and mutual adjustment | 1.22<br>(0.82, 1.82) | 1.44<br>(0.94, 2.21) |  |

\*RMSE for 2- and 3-way models was calculated as follows: $RMSE = \sqrt{\sum (P_i - O_i)^2 / n}$ . \*\*According to MV + mutual adjustment. Bold values denote $p < 0.05$

**Table 4:** Multi-omics inflammation signature for TNFa and association with all-cause mortality in CLSA

| N omics | Approach | The number and type of predictors included in the final model | Sample size to calculate model performance | RMSE* | R (Pearson) | Analysis model | 1-SD of blood CRP | 1-SD of the signature | Sample size and number of events** |
| --- | --- | --- | --- | --- | --- | --- | --- | --- | --- |
| 3-way: PRS-MRS-ERS | Hierarchical | TNFa predicted by 1 PRS, TNFa predicted by 110 CpGs, TNFa predicted by 134 metabolites | 1,392 | 0.130 | 0.574 | Age, sex, smoking, multimorbidity, BMI, race/ethnicity, and alcohol-adjusted model (MV) | **1.62**<br>**(1.22, 2.16)** | 1.12<br>(0.82, 1.54) | N=917; number of events=44 |
|  |  |  |  |  |  | MV + and mutual adjustment | **1.61**<br>**(1.20, 2.16)** | 1.04<br>(0.75, 1.44) |  |
| 2-way: PRS-MRS | Hierarchical | 1 PRS + predicted value by 134 metabolites | 9,357<br>[1,392] | 0.124<br>[0.132] | 0.607<br>[0.569] | Age, sex, smoking, multimorbidity, BMI, race/ethnicity, and alcohol-adjusted model (MV) | **1.24**<br>**(1.09, 1.41)** | **1.32**<br>**(1.21, 1.44)** | N=6272; number of events=266 |
|  |  |  |  |  |  | MV + and mutual adjustment | 1.05<br>(0.91, 1.21) | **1.30**<br>**(1.18, 1.44)** |  |
| 2-way: PRS-ERS | Hierarchical | 1 PRS + 139 CpGs | 1,393 | 0.145 | 0.413 | Age, sex, smoking, multimorbidity, BMI, race/ethnicity, and alcohol-adjusted model (MV) | **1.68**<br>**(1.28, 2.20)** | **1.72**<br>**(1.19, 2.48)** | N=918; number of events=44 |
|  |  |  |  |  |  | MV + and mutual adjustment | **1.50**<br>**(1.09, 2.07)** | **1.46**<br>**(1.00, 2.13)** |  |
| 2-way: MRS-ERS | Hierarchical | predicted value by 50 CpGs and predicted value by 261 metabolites | 1,425 | 0.129 | 0.577 | Age, sex, smoking, multimorbidity, BMI, race/ethnicity, and alcohol-adjusted model (MV) | **1.69**<br>**(1.28, 2.23)** | **1.90**<br>**(1.43, 2.52)** | N=937; number of events=46 |
|  |  |  |  |  |  | MV + and mutual adjustment | 1.31<br>(0.93, 1.83) | **1.66**<br>**(1.19, 2.33)** |  |
| 1-way: PRS | PRS-CS | 1 PRS | 9,360 | 0.273 | 0.032 | Age, sex, smoking, multimorbidity, BMI, race/ethnicity, and alcohol-adjusted model (MV) | **1.22**<br>**(1.07, 1.39)** | 1.02<br>(0.90, 1.14) | N=6275; number of events=266 |
|  |  |  |  |  |  | MV + and mutual adjustment | **1.22**<br>**(1.07, 1.39)** | 1.02<br>(0.90, 1.15) |  |
| 1-way: MRS | 10-fold CV elastic net | 261 metabolites | 9,554 | 0.123 | 0.610 | Age, sex, smoking, multimorbidity, BMI, race/ethnicity, and alcohol-adjusted model (MV) | **1.24**<br>**(1.09, 1.41)** | **1.27**<br>**(1.19, 1.36)** | N=6412; number of events=275 |
|  |  |  |  |  |  | MV + and mutual adjustment | 1.09<br>(0.95, 1.24) | **1.25**<br>**(1.17, 1.36)** |  |
| 1-way: ERS | 10-fold CV elastic net | 363 CpGs | 1,426 | 0.146 | 0.376 | Age, sex, smoking, multimorbidity, BMI, race/ethnicity, and alcohol-adjusted model (MV) | **1.69**<br>**(1.28, 2.23)** | **1.48**<br>**(1.17, 1.87)** | N=938; number of events=46 |
|  |  |  |  |  |  | MV + and mutual adjustment | **1.60**<br>**(1.19, 2.15)** | **1.37**<br>**(1.05, 1.77)** |  |

\*RMSE for 2- and 3-way models was calculated as follows: $RMSE = \sqrt{\sum (P_i - O_i)^2 / n}$ . \*\*According to MV + mutual adjustment. Bold values denote $p < 0.05$

**Table 5:** Validation of the CRP and IL6 risk scores in NHS I, NHS II, and HPFS

|  | NHS I |  |  | NHS II |  |  | HPFS |  |  |
| --- | --- | --- | --- | --- | --- | --- | --- | --- | --- |
|  | R<br>(Pearson) | P | N | R<br>(Pearson) | P | N | R<br>(Pearson) | P | N |
| **CRP** |  |  |  |  |  |  |  |  |  |
| ERS* - blood draw #1 | - | - | - | 0.329 | <0.001 | 334 | - | - | - |
| ERS* – blood draw #2 | - | - | - | 0.175 | 0.004 | 253 | - | - | - |
| MRS** | 0.371 | <0.001 | 4321 | 0.445 | <0.001 | 580 | 0.194 | <0.001 | 725 |
| PRS | 0.440 | <0.001 | 8,409 | 0.405 | <0.001 | 1,736 | 0.414 | <0.001 | 3,887 |
| MRS-ERS | - | - | - | 0.312 | 0.042 | 43 | - | - | - |
| PRS-MRS | 0.555 | <0.001 | 2,412 | 0.507 | <0.001 | 180 | 0.415 | <0.001 | 506 |
| PRS-ERS | - | - | - | 0.523 | <0.001 | 253 | - | - | - |
| PRS-MRS-ERS | - | - | - | 0.416 | 0.01 | 37 | - | - | - |
| **IL6** |  |  |  |  |  |  |  |  |  |
| ERS* - blood draw #1 | - | - | - | 0.313 | <0.001 | 135 | - | - | - |
| ERS* – blood draw #2 | - | - | - | 0.257 | 0.007 | 51 | - | - | - |
| MRS** | 0.312 | <0.001 | 3,333 | 0.343 | <0.001 | 696 | 0.238 | <0.001 | 813 |
| PRS | 0.698 | <0.001 | 5,181 | 0.703 | <0.001 | 1,428 | 0.691 | <0.01 | 2,546 |
| MRS-ERS | - | - | - | 0.385 | 0.16 | 15 | - | - | - |
| PRS-MRS | 0.460 | <0.001 | 1,953 | 0.529 | <0.001 | 187 | 0.440 | <0.001 | 619 |
| PRS-ERS | - | - | - | 0.428 | <0.001 | 89 | - | - | - |
| PRS-MRS-ERS | - | - | - | 0.597 | 0.031 | 13 | - | - | - |

\*7 CpGs removed from the original 150-CpG ERS model due to differences in DNA methylation QC (removing probes)
\*\*234 metabolites were removed from NHS, NHS II, and HPFS datasets due to the differences in the metabolomic platforms (blank HMDB).

#### Inflammation MRS

Our MRS generated using 10-fold CV resulted in a prediction of the blood markers by 188-400 (CRP, N=9,956, R=0.688), 165-305 (IL6, N=9,727, R=0.656), and 95-261 (TNFa, N=9,554, R=0.610) metabolites (**Tables 2-4, Figure S3**). Next, we validated our MRS for CRP and IL6 in the NHS, NHS II, and HPFS data using our final model with 319 and 428 metabolites for CRP and IL6 risk scores. The MRS for CRP performed well in the NHS, NHS II, and HPFS (**Table 5**). Of note, 234 metabolites were removed from NHS, NHS II, and HPFS datasets (**Table S2**) due to the differences in the metabolomic platforms (blank HMDB). The MRS for IL6 showed similar performance across all validation cohorts (**Table 5**).

#### Inflammation ERS

Our ERS generated using 10-fold CV resulted in a prediction of the blood markers by 122-328 (CRP, N=1,468, R=0.534), 356-527 (IL6, N=1,418, R=0.575), and 137 to 769 (TNFa, N=1,426, R=0.376) CpGs (**Tables 2-4, Figure S4a-c**). We also applied previously published epigenetic scores to predict CRP and IL6 in the CLSA data and correlated these with the actual blood levels of each marker. This resulted in a correlation of R=0.183 and R=0.256 for blood CRP vs. the published Ligthart and Barker CRP epigenetic scores and R=0.123 for blood IL6 vs. published Stevenson IL6 epigenetic score.

Validation of ERS for CRP and IL6 using the NHS II data showed a good correlation between the ERS and the blood levels of each marker (CRP: R=0.329 and R=0.175, IL6: R=0.313 and R=0.257; examined in two blood draws in the NHS II study; **Table 5**).

### Two-way omics risk scores for inflammation markers

Of the three 2-way integrated risk scores (PRS-MRS, PRS-ERS, or MRS-ERS), the PRS-MRS for CRP, IL6, and TNFa had the highest correlation with the corresponding blood markers (**Tables 2-4**). To examine if this observation is due to the different available samples for these risk scores, we repeated the correlation test in a subset using the same set of samples as further used for the 3-way signatures. For CRP, the PRS-MRS remained the strongest of the 2-way risk score, but for IL6 and TNFa, the 2-way MRS-ERS had the strongest correlation with the blood markers of all the 2-way scores. Validation of the 2-way PRS-MRS (**Table 5**) showed a good correlation between the signature and blood levels of CRP and IL6. The 2-way PRS-ERS and MRS-ERS showed significant correlation between the scores and corresponding blood levels of the markers in the NHS II (**Table 5**), except for IL6 MRS-ERS.

### Three-way scores based on PRS, MRS, and ERS

The fully integrated (3-way) scores included the PRS, MRS, and ERS. When considering the same sample size and the same set of samples used to calculate the 2-way risk-score subset correlation test, the 3-way CRP, IL6, and TNFa risk scores showed the strongest correlation with the corresponding blood markers (**Tables 2-4**). All models (1-2-3-way risk scores for all three inflammation markers) are presented in **Tables S3-TableS7**. The 3-way CRP and IL6 risk scores performed well in the NHS II validation set (**Table 5**), though the sample sizes were limited (N=37 for CRP and N=13 for IL6).

We repeated the scorers’ validation in the NHS, NHS II, and HPFS using a minimal sample size that included only individuals with both CRP and IL6 measurements instead of all available sample size per inflammation marker and omics (**Table S8**). This validation yielded similar results.

#### Blood CRP and CRP omics risk scores as predictors for all-cause mortality

We performed a series of multivariate models, detailed in **Table S9**. When adjusting for age, sex, smoking, multimorbidity, BMI, race/ethnicity, and alcohol (**Table 2**), the MRS-ERS for CRP (N=1,430) had the highest HR (1.76, 95% CI [1.22, 2.53]). In mutually adjusted models, considering both blood CRP and each omics CRP signature in the same Cox model detailed above, only the MRS-ERS, MRS, and ERS remained significant predictors for all-cause mortality. The 2-way MRS-ERS remained the strongest predictor for all-cause mortality, compared with the other 2-way risk scores (HR=1.77, 95% CI [1.14,2.77]). When input in a mutually adjusted model with blood CRP, the 3-way CRP risk score was not a significant predictor for all-cause mortality.

We performed a series of 1-df LRTs, examining the above models with and without the CRP risk scores (e.g., multivariate models including the blood marker – “mutual adjustment”, with and without 1-way ERS, with and without 2-way PRS-ERS, etc.,). For a total of 7 LRTs, one for each CRP risk score, the following LRTs were significant: ERS, MRS, and MRS-ERS (p<0.05 for all). CRP PRS, PRS-MRS, and PRS-MRS-ERS LRTs were marginally significant (p=0.09, p=0.07, and p=0.06, respectively).

#### Blood IL6 and IL6 omics risk scores as predictors for all-cause mortality

We repeated the same models to test for association with all-cause mortality, with either blood IL6, different omics risk scores for IL6 (N=1,384), or a mutual adjustment (**Table 3**). Full models details, coefficients, and p-values are presented in **Table S10**. The 3-way IL6 risk scores and the 2-way MRS-ERS had the highest HR of all mutually adjusted models (HR=1.62, 95% CI [1.04,2.53] and HR=1.77, 95% CI [1.15,2.72], respectively), while the blood IL6 was not a significant predictor for all-cause mortality in these models. In 1-df LRTs, examining the above models of mutual adjustment with and without the IL6 risk scores, the following LRTs were significant: MRS, PRS-MRS, MRS-ERS, and PRS-MRS-ERS (p<0.05 for all). 1-df LRT for IL6 ERS was marginal (p=0.09).

#### Blood TNFa and TNFa omics risk scores as predictors for all-cause mortality

Finally, we repeated the same Cox regression models with either blood TNFa (N=1,392), different risk scores for TNFa, or a mutual adjustment (**Table 4**). Full models details, coefficients, and p-values are presented in **Table S11**. The MRS, ERS, and all 2-way risk scores were significantly associated with all-cause mortality in the mutually adjusted models (MRS: HR:1.25 [1.17,1.36]; ERS: 1.37 [1.05,1.77]; MRS-ERS: HR=1.66 [1,19,2.33]; PRS-ERS: HR=1.46 [1.00,2.13]; PRS-MRS: HR=1.30 [1.18,1.44]). In 1-df LRTs, examining the above models of mutual adjustment with and without the TNFa risk scores, all LRTs were significant (p<0.05 for all), except for the TNFa PRS and PRS-MRS-ERS LRTs (p=0.78 and p=0.81).

#### Sensitivity analysis and prediction improvement by omics

A series of sensitivity analyses (**Table S12-S19**) using logistic regression to get the Odds Ratio in comparable models to the survival analysis yielded similar conclusions.

Next, we first repeated the Cox analyses while inputting first the traditional predictors (**model 1**: age, sex, smoking; **model 2**: model 1 + BMI; **model 3**: model 2 + race/ethnicity; **model 4:** model 3 + alcohol intake; **model 5**: model 4 + multi-morbidity). We then included omics risk scores (**model 6**: model 5 + the 3-way risk scores for either CRP, IL6, or TNFa) and examined the change in C-statistics as an indicator for model improvement. The change in concordance for the three 3-way risk scores is presented in **Figure 2**. Examining the dynamic prediction of model 6 using the AUC showed that the full model with the 3-way TNFa risk score had the highest AUC, followed by the IL6 and the CRP scores (AUC= 0.789, 0.772, 0.744; TNFa, IL6, and CRP, respectively; **Figure S5**). Inputting either of the 3-way risk scores resulted in the highest concordance, compared with models with traditional markers for mortality. Adding all three 3-way risk scores to model 5 resulted in an AUC of 0.805.

**Figure 2(a-c):**
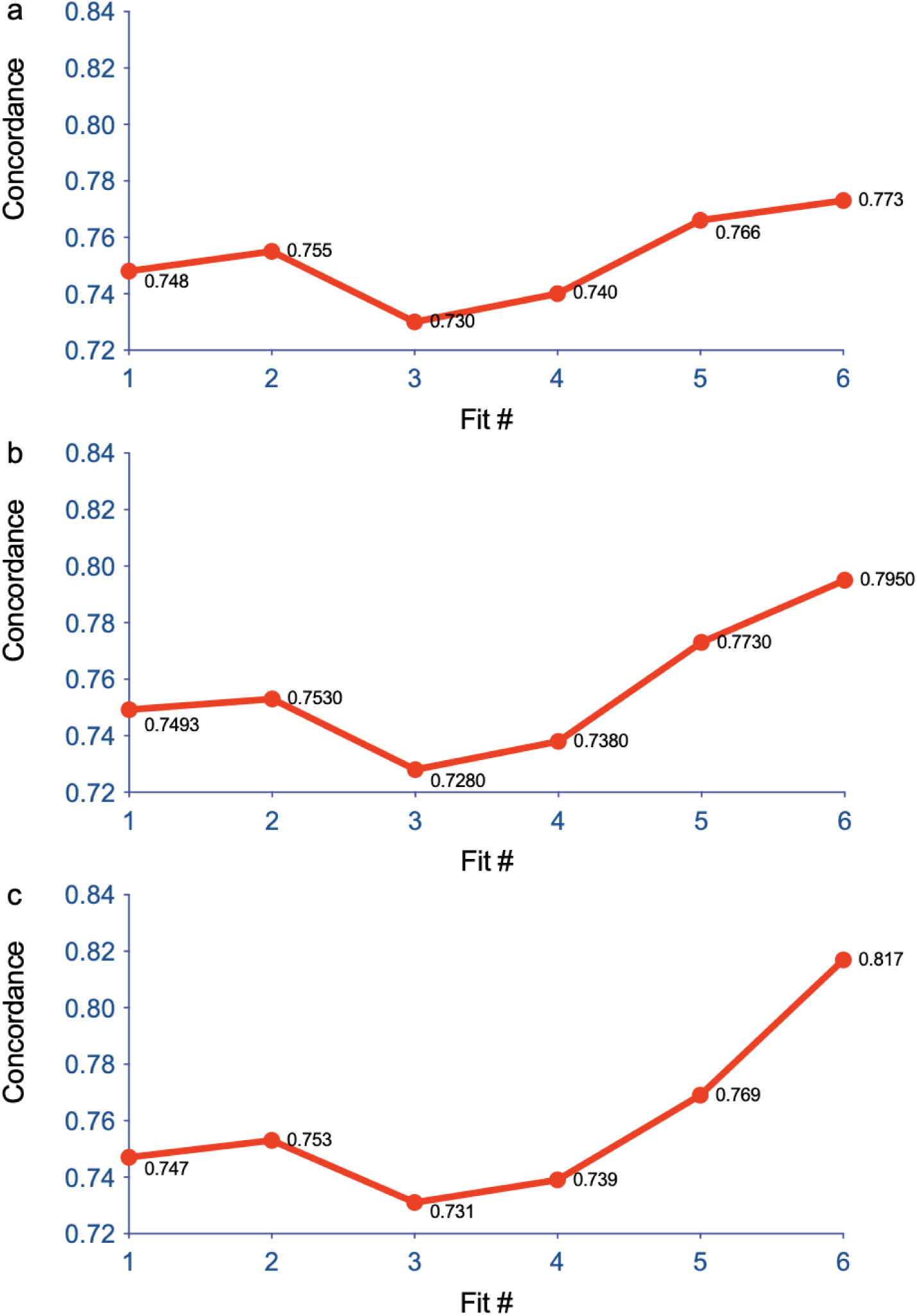
Changes in concordance across different models (fit). (a) CRP 3-way risk score input in the last model (fit6); (b) IL6 3-way risk score input in the last model (fit6); (c) TNFa 3-way risk score input in the last model (fit6). List of models: Model 1 (fit1): age, sex, and smoking. Model 2 (fit2): age, sex, smoking, and BMI. Model 3 (fit3): age, sex, smoking, BMI, and race/ethnicity. Model 4 (fit4): age, sex, smoking, BMI, race/ethnicity, and alcohol intake. Model 5 (fit5): age, sex, smoking, BMI, race/ethnicity, alcohol intake, multi-morbidity. Model 6 (fit6): age, sex, smoking, BMI, race/ethnicity, alcohol intake, and multi-morbidity, and the 3-way risk score (a: CRP; b: IL6; c: TNFa).

#### The joint effect of the CRP, IL6, and TNFa omics risk scores on all-cause mortality

As each blood inflammation marker may represent a different aspect of inflammation, we included all 3-way risk scores and all blood markers for the subsequent analysis. We followed the same models detailed above (**Table S19**). In a model including only the three 3-way risk scores and blood biomarkers to predict all-cause mortality, only the TNFa and TNFa risk score were significantly associated with all-cause mortality (HR=1.43 [1.07, 1,91] and HR=1.48 [1.12, 1.97] blood and risk score, respectively). Yet, when further adjusted for confounders, none of the risk scores or blood markers remained significantly associated with all-cause mortality. We continued to explore the joint effect of the risk scores using a LRT test. In a model including all three 3-way risk scores, blood markers, age, sex, smoking, multimorbidity, BMI, race/ethnicity, and alcohol intake, the 3-df LRT was insignificant after removing all risk scores (p=0.10).

### Validation of the associations with all-cause mortality

Finally, we sought to validate the results of the association of the risk scores with all-cause mortality in the validation cohorts. The median follow-up time was 28.0, 21.0, and 20.1 years, NHS, NHS II, and HPFS, respectively. We combined these cohorts due to a smaller sample size with overlapping omics between participants measured allowed us to use the following risk scores in addition to blood CRP and IL6: PRS, MRS, and PRS-MRS. We used the following comparable stepwise models to examine the HR between the CLSA, NHS, NHS II, and HPFS (by the available omics): age + sex + smoking + BMI + race/ethnicity + alcohol intake (full model; **Table S20-21**). CLSA and NHS/HPFS showed the same direction of effect size to the association between blood CRP and blood IL6 and mortality. Similarly, for the MRSs for IL6 and CRP, most models’ results were similar between the cohorts, with a stronger effect size in the CLSA. The IL6 2-way MRS-PRS showed the same direction in all cohorts, but the magnitudes differed between the CLSA and the NHS/HPFS.

To better align the average age of the baseline samples in CLSA and NHS/HPFS, we further restricted the models to participants age > 65 years in CLSA (N=12,646) and NHS/HPFS (N=2,020; **Table S22-23**); this slightly increased the HR observed in the NHS/HPFS across the different risk scores and blood markers.

## Discussion

In this study, we established single and multi-omics risk scores to evaluate blood inflammation markers. We validated these risk scores in three cohorts and compared our ERS to previously published scores, showing the increased predictive ability for mortality.

We designed a “hierarchical approach” to build our multi-omics models sequentially by leveraging the power of different omics’ available sample size and maximizing the residual total variance explained by subsequence omics risk score. At each step, we used a 10-fold CV to ensure the model used to compute the predicted score for each subject is completely independent of the subject in the calculation. Instead of inputting all coefficients from the 1-way scores to build a multi-omics score, this approach helps avoid overfitting the models, maximizes the variance explained by all involved omics predictors, and allows exploiting the changing sample size available per omics. Compared with previously published ERS for CRP built on Epigenome Wide Association Study results and 450K methylation array data (Ligthart ^25^ and Barker ^26^), our CRP ERS model outperformed the published ones with a higher correlation between the CRP ERS and blood CRP. This can be explained by using penalized regression for candidate predictors rather than performing EWAS, which reduces overfitting and provides the combined effect of predictors to get the highest explained variance, rather than single predictors ^28^. Also, additional CpGs in the EPIC array compared with the 450K array may possess additional biological information critical to evaluating inflammatory state. Our ERS for IL6 also outperformed the previously published score by Stevenson ^27^, although both methods relied on penalized regression to establish the DNA methylation IL6 score. Yet, Stevenson et al. methylation data was subset to probes in common on both the 450k and EPIC arrays to establish cross-array scores, as their training sample (Lothian Birth Cohort) used 450K array to assess DNA methylation and applied the IL6 score to the Generation Scotland with DNA methylation analyzed by the EPIC array.

The omics risk scores that included metabolomics showed the highest prediction of blood inflammation markers. It has been suggested that metabolites from different tissues are involved in regulating the activation of the immune system and, thus, predict inflammatory-based diseases’ outcomes ^29^. A previous study used blood metabolomics from three biobanks to build MRS to identify high-risk groups for 12 diseases ^30^. In that study, the MRS was a stronger predictor for most future disease onset than PRS. That study also suggested that a joint PRS-MRS model may be the best-performing model for disease prediction. Integrating different omics from different functional layers, some subjected to external exposures such as metabolomics and epigenetics, and genetics, representing the inheritable risk of diseases, may provide complementary biological information on a phenotype or capture the confounding effect of other correlated phenotypes/exposures. This was supported by our results, with an increase in the performance of the prediction models, from single to multi-omics risk scores.

Our central hypothesis was that integrating several time-scope omics from different function layers may provide additional biological potential on mortality, not yet reflected in the blood levels of the biomarkers measured at the same blood draw. When associated with mortality and compared to the actual blood levels of each marker, the risk scores outperformed the blood markers in terms of effect size and significance in most models, compared to CRP and IL6, but not for TNFa. Additionally, when examining the contribution of each score to the association with all-cause mortality, by including all covariates starting with the traditional background factors and last for the omics risk scores, we found that either of the 3-way risk scores resulted in the highest concordance, compared with models with traditional factors for mortality (age, sex, smoking, BMI, race/ethnicity, alcohol intake, multi-morbidity). An elevated inflammatory profile reflecting chronic low-grade inflammation was shown to be a predictor for all-cause mortality. A previous meta-analysis of 14 studies showed that elevated blood CRP is an independent risk factor for all-cause mortality in the general population ^6^. This was also demonstrated in middle-aged and older men and women above 65 years ^9^. IL6 predicted all-cause mortality among elderly men over 65 years ^8^. Other studies identified blood IL6 and TNFa as predictors for all-cause mortality, mostly among older population and with chronic diseases ^8,31,32^. Among the general population in our study, some biological risk scores, such as 3-way IL6 risk score, 2-way CRP MRS-ERS, and 2-way TNFa MRS-ERS, were found to be stronger predictors for all-cause mortality, compared with blood inflammation levels. When trying to validate the results of the association study between omics inflammation markers, blood markers, and all-cause mortality using the NHSs and HPFS, results were consistent between the cohorts, with smaller but similar directions of effect size for IL6 risk scores and blood IL6 and CRP, but inconsistent for CRP risk scores and mutual adjustment models. These can be explained by the smaller sample size in the NHS/HPFS, younger age compared to the CLSA, and potential environmental exposures between the residential areas where all participants live.

Our study has several limitations. First, we could not externally validate TNFa models since TNFa was not measured in any of the NHS/HPFS datasets. Second, the CLSA and NHS/HPFS used different metabolomic platforms and had fewer overlapping metabolites than the number of metabolites used in training the models (CLSA). Third, only one validation cohort (NHS II) had DNA methylation measured and was used to validate the 3-way risk score. Although this cohort had a substantially smaller sample size, we observed good performance for our 3-way CRP and IL6 risk scores. Forth, some of the associations with all-cause mortality were not statistically significant in the NHS and HPFS, probably due to limited sample size and the smaller number of overlapping metabolomics all datasets, a larger percentage of women in the validation set, the timing in terms of participant age of measuring the metabolomics, the mean age at the beginning of the follow-up (stratifying on >65 years old subjects improved the validation performance), the duration of the follow-up, and differences in mortality rates due to different geographical environment. Nevertheless, our study has strengths to be highlighted. We integrated three omics to produce 1-2- and 3-way risk scores for inflammation markers. Our scores out-performed previously published ERS. We demonstrated that including at least 2 omics as predictors of inflammation markers improves the model’s predictive ability.

In conclusion, we have developed multi-omics inflammation signature models to capture burden of inflammation at a more comprehensive spectrum and different time-scores. Utilizing omics as risk scores of inflammation predicted all-cause mortality beyond the snapshot of blood traditional biomarkers, age, sex, and other background characteristics, including morbidity. This may provide a method to identify population subject to inflammation-based morbidity and mortality for better target intervention.

## Acknowledgments

CLSA: This research was made possible using the data/biospecimens collected by the Canadian Longitudinal Study on Aging (CLSA). This research has been conducted using the CLSA Baseline Comprehensive Dataset version CoPv7, comprehensive Follow-up 1 (version 3.2), Dataset A Participant Status (version 2.0), CLSA Genome-wide genetic data (version 3.0), CLSA DNA Methylation data (version 1.1), and CLSA Metabolomics data (version 1.0) under Application ID 2109001. The CLSA is led by Drs. Parminder Raina, Christina Wolfson and Susan Kirkland. The opinions expressed in this manuscript are the author’s own and do not reflect the views of the Canadian Longitudinal Study on Aging.

NHS, NHS II, and HPFS: The authors would like to acknowledge the contribution to this study from central cancer registries supported through the Centers for Disease Control and Prevention’s National Program of Cancer Registries (NPCR) and/or the National Cancer Institute’s Surveillance, Epidemiology, and End Results (SEER) Program. Central registries may also be supported by state agencies, universities, and cancer centers. Participating central cancer registries include the following: Alabama, Alaska, Arizona, Arkansas, California, Colorado, Connecticut, Delaware, Florida, Georgia, Hawaii, Idaho, Indiana, Iowa, Kentucky, Louisiana, Massachusetts, Maine, Maryland, Michigan, Mississippi, Montana, Nebraska, Nevada, New Hampshire, New Jersey, New Mexico, New York, North Carolina, North Dakota, Ohio, Oklahoma, Oregon, Pennsylvania, Puerto Rico, Rhode Island, Seattle SEER Registry, South Carolina, Tennessee, Texas, Utah, Virginia, West Virginia, Wyoming.

## Data Availability statement

Data are available from the Canadian Longitudinal Study on Aging (www.clsa-elcv.ca) for researchers who meet the criteria for access to de-identified CLSA data.

Further information including the procedures to obtain and access data from the Nurses’ Health Studies and Health Professionals Follow-up Study is described at https://www.nurseshealthstudy.org/researchers (contact) and https://sites.sph.harvard.edu/hpfs/for-collaborators/.

## Ethics

CLSA: Ethical review of the CLSA protocol was conducted by the research ethics board at each research site with the coordination of the McMaster Research Ethics Board (at baseline there were 13 REBs involved).

NHS, NHS II, HPFS: The study protocol was approved by the institutional review boards of the Brigham and Women’s Hospital and Harvard T.H. Chan School of Public Health, and those of participating registries as required.

## References

1. Aul, P. et al. C-Reactive Protein and Other Markers of Inflammation in the Prediction of Cardiovascular Disease in Women. 10.1056/NEJM2000032334212021, 1066–1067 (2000).

2. Zhang, W. et al. High-Sensitivity C-Reactive Protein Modifies the Cardiovascular Risk of Lipoprotein(a): Multi-Ethnic Study of Atherosclerosis. J Am Coll Cardiol 78, 1083–1094 (2021).

3. Okdahl, T. et al. Low-grade inflammation in type 2 diabetes: A cross-sectional study from a Danish diabetes outpatient clinic. BMJ Open 12, e062188 (2022).

4. Kinney, J. W. et al. Inflammation as a central mechanism in Alzheimer’s disease. Alzheimer’s & Dementia: Translational Research & Clinical Interventions 4, 575–590 (2018).

5. Franceschi, C., Garagnani, P., Parini, P., Giuliani, C. & Santoro, A. Inflammaging: a new immune–metabolic viewpoint for age-related diseases. Nat Rev Endocrinol 14, 576–590 (2018).

6. Li, Y. et al. Hs-CRP and all-cause, cardiovascular, and cancer mortality risk: a meta-analysis. Atherosclerosis 259, 75–82 (2017).

7. Brüünsgaard, H. & Pedersen, B. K. Age-related inflammatory cytokines and disease. Immunology and Allergy Clinics 23, 15–39 (2003).

8. Baune, B. T., Rothermundt, M., Ladwig, K. H., Meisinger, C. & Berger, K. Systemic inflammation (Interleukin 6) predicts all-cause mortality in men: results from a 9-year follow-up of the MEMO Study. Age (Omaha) 33, 209–217 (2011).

9. Li, Z.-H. et al. Associations of plasma high-sensitivity C-reactive protein concentrations with all-cause and cause-specific mortality among middle-aged and elderly individuals. Immunity & Ageing 16, 1–8 (2019).

10. Raina, P. et al. Cohort profile: the Canadian longitudinal study on aging (CLSA). Int J Epidemiol 48, 1752–1753j (2019).

11. Forgetta, V. et al. Cohort profile: genomic data for 26 622 individuals from the Canadian Longitudinal Study on Aging (CLSA). BMJ Open 12, e059021 (2022).

12. Forgetta, V. et al. The Canadian Longitudinal Study on Aging Genome-Wide Genetic Data Release (Version 3). (2020).

13. UKB_WCSGAX: UK Biobank 500K Samples Genotyping Data Generation by the Affymetrix1 Research Services Laboratory. appliedbiosystems https://biobank.ctsu.ox.ac.uk/crystal/crystal/docs/affy_data_generation2017.pdf (2007).

14. A reference panel of 64,976 haplotypes for genotype imputation. Nat Genet 48, 1279–1283 (2016).

15. Michelotti, G. et al. Metabolomic Profiling on 9,992 Participants Using Ultra-Performance Liquid Chromatography and Mass Spectrometer Data Support Document. (2023).

16. Tse Shen Lin, D., et al. Genome-Wide DNA Methylation Profiling on 1,478 Participants Using Illumina Infinium MethylationEPIC BeadChip Microarray Technology Data Support Document. (2022).

17. Verschoor, C. P. et al. Epigenetic age is associated with baseline and 3-year change in frailty in the Canadian Longitudinal Study on Aging. Clin Epigenetics 13, 1–10 (2021).

18. Aryee, M. J. et al. Minfi: a flexible and comprehensive Bioconductor package for the analysis of Infinium DNA methylation microarrays. Bioinformatics 30, 1363–1369 (2014).

19. Verschoor, C. P., Vlasschaert, C., Rauh, M. J. & Paré, G. A DNA methylation based measure outperforms circulating CRP as a marker of chronic inflammation and partly reflects the monocytic response to long-term inflammatory exposure: A Canadian longitudinal study of aging analysis. Aging Cell e13863 (2023).

20. Pietzner, M. et al. Plasma metabolites to profile pathways in noncommunicable disease multimorbidity. Nat Med 27, 471–479 (2021).

21. Sun, Q. et al. Physical Activity at Midlife in Relation to Successful Survival in Women at Age 70 Years or Older. Arch Intern Med 170, 194–201 (2010).

22. Petersen, L. K. et al. Understanding and Predicting Polycystic Ovary Syndrome through Shared Genetics with Testosterone, SHBG, and Chronic Inflammation. medRxiv 2010– 2023 (2023).

23. Willer, C. J., Li, Y. & Abecasis, G. R. METAL: fast and efficient meta-analysis of genomewide association scans. Bioinformatics 26, 2190–2191 (2010).

24. Wainberg, M. et al. Genetic architecture of the structural connectome. Nat Commun 15, 1–20 (2024).

25. Ligthart, S. et al. DNA methylation signatures of chronic low-grade inflammation are associated with complex diseases. Genome Biol 17, 1–15 (2016).

26. Barker, E. D. et al. Inflammation-related epigenetic risk and child and adolescent mental health: A prospective study from pregnancy to middle adolescence. Dev Psychopathol 30, 1145–1156 (2018).

27. Stevenson, A. J. et al. Creating and validating a DNA methylation-based proxy for interleukin-6. The Journals of Gerontology: Series A 76, 2284–2292 (2021).

28. Yousefi, P. D. et al. DNA methylation-based predictors of health: applications and statistical considerations. Nat Rev Genet 23, 369–383 (2022).

29. Fitzpatrick, M. & Young, S. P. Metabolomics–a novel window into inflammatory disease. Swiss Med Wkly 143, w13743 (2013).

30. Group, N. H. B. C. et al. Metabolomic and genomic prediction of common diseases in 477,706 participants in three national biobanks. medRxiv 2023–2026 (2023).

31. Sun, J. et al. Biomarkers of cardiovascular disease and mortality risk in patients with advanced CKD. Clin J Am Soc Nephrol 11, 1163 (2016).

32. Tripepi, G., Mallamaci, F. & Zoccali, C. Inflammation markers, adhesion molecules, and all-cause and cardiovascular mortality in patients with ESRD: searching for the best risk marker by multivariate modeling. Journal of the American Society of Nephrology 16, S83– S88 (2005).

