## Supplemental material for "Cross omics risk scores of inflammation markers are associated with all-cause mortality: The Canadian Longitudinal Study on Aging"

Anat Yaskolka Meir^1^*, Huan Yun^1^, Jie Hu^1,2^, Jun Li^3^, Jiaxuan Liu^1^, Alaina Bever^1^, Andrew Ratanatharathorn^1^, Mingyang Song^1,4,5,6^, A. Heather Eliassen^1,7,8^, Lori Chibnik^1^, Karestan Koenen^1^, Guillaume Pare^9^, Meir J Stampfer^1,7,8^, Liming Liang^1,10^*

*Corresponding authors

^1^ Department of Epidemiology, Harvard T.H. Chan School of Public Health, Boston, MA 02115, USA

^2^ Center for Genomic Medicine and Department of Anesthesia, Critical Care and Pain Medicine, Massachusetts General Hospital and Harvard Medical School, Boston, MA 02114, USA

^3^ Division of Preventive Medicine, Department of Medicine, Brigham and Women’s Hospital and, Harvard Medical School, Boston, MA 02115, USA

^4^ Clinical and Translational Epidemiology Unit, Massachusetts General Hospital and Harvard Medical School, Boston, MA, USA.

^5^ Division of Gastroenterology, Massachusetts General Hospital and Harvard Medical School, Boston, MA, USA.

^6^ The Broad Institute of MIT and Harvard, Cambridge, MA, USA.

^7^ Department of Nutrition, Harvard T.H. Chan School of Public Health, Boston, MA 02115, USA

^8^ Channing Division of Network Medicine, Department of Medicine, Brigham & Women’s Hospital & Harvard Medical School, Boston, MA 02115, USA

^9^ Department of Health Research Methods, Evidence and Impact, McMaster University, Hamilton, Ontario, Canada

^10^ Department of Biostatistics, Harvard T.H. Chan School of Public Health, Boston, MA 02115, USA

Correspondence

**Dr. Liming Liang**

Department of Epidemiology, Harvard T.H. Chan School of Public Health. 655 Huntington Avenue, Boston, MA 02115, USA.

**Dr. Anat Yaskolka Meir**

Department of Epidemiology, Harvard T.H. Chan School of Public Health. 655 Huntington Avenue, Boston, MA 02115, USA.

**Methods S1: description of the validation cohorts**

The Nurse’s Health Study (NHS) was initiated in 1976 and included 121,700 female nurses aged 30–55 years ^1^. The NHSII was initiated in 1989 and included 116,429 female nurses aged 25–42 years ^1^. The Health Professional Follow-up Study (HPFS) was initiated in 1986 and included 51,529 male health professionals aged 40–75 years ^2^. The baseline questionnaire included information regarding lifestyle, medical history, and health-related questions and was followed up biennially. The study protocol was approved by the institutional review boards of the Brigham and Women’s Hospital and Harvard T.H. Chan School of Public Health, and those of participating registries as required.

**Methods S2: Omics in the validation cohorts**

Omics data collection and preprocessing were detailed before for genetic and blood metabolomic ^3^. Briefly: Genotyping in NHS, NHS II, and HPFS was performed using five whole-genome arrays and was merged and imputed based on the Haplotype Reference Consortium reference panel. Genetic variants with a minor allele frequency >1% and imputation quality >0.3 were used in analyses.

Genomic DNA was extracted in the NHS II only using the QIAamp 96-spin blood protocol (Qiagen, Valencia, California).

The plasma metabolomics profiling for the NHS, NHS II, and HPFS were at the Broad Institute of Harvard University and M.I.T. (Cambridge, MA), using high-throughput liquid chromatography-tandem mass spectrometry techniques. After quality filtering and standardization, a total of 396 named metabolites were qualified in the NHS/HPFS.

DNAm was assessed using the Illumina HumanMethylation EPIC BeadArray. Data was processed using a quality control pipeline laid out by the Psychiatrics Genomics Consortium’s PTSD Epigenetics working group: <https://github.com/PGC-PTSD-EWAS/EPIC_QC>), including Illumina control metrics check, normalization, exclusion of low-quality samples and probes, and exclusion of potentially cross-hybridizing probes. A total of 1,538 samples (two time points) and 819,998 CpGs on autosomal and X chromosomes passed quality control.

**Methods S3: Measurement of blood inflammation markers in the validation cohorts**

Blood samples to measure CRP and IL6 were collected from subsamples of the NHS during 1989–1990, NHS II during 1996–1999 & 2010-2012, and HPFS during 1993–1995 (follow-up rate >95%). The Institutional Review Boards at Brigham and Women’s Hospital and Harvard T.H. Chan School of Public Health approved the study. Further details on blood collection and measurements can be found elsewhere ^4^.

**Table S1: Background characteristics of the validation sets across the availability of omics data and corresponding blood markers^1^**

| Characteristics | NHS, NHSII, HPFS studies: individuals with genetic data | | NHS, NHSII, HPFS studies: individuals with metabolomic data | | NHS II: individuals with epigenetic data | |
| --- | --- | --- | --- | --- | --- | --- |
| Minimal and maximal sample size used for 1-way signatures | Minimal sample size used for PRS (N=9,155) | Maximal sample size used for PRS (N=14,032) | Minimal sample size used for MRS (N=4,842) | Maximal sample size used for MRS (N=5,626) | Minimal sample size used for ERS (N=51) | Maximal sample size used for ERS (N=334) |
| Age | 56.15(9.70) | 57.00(9.31) | 56.45(8.81) | 57.12(8.31) | 44.04(4.68) | 44.45(4.60) |
| Men, % | 1725(18.8%) | 2632(18.8%) | 536(11.1%) | 457(8.1%) | 0 | 0 |
| Body mass index | 26.0(5.02) | 26.15(5.04) | 25.86(4.81) | 25.98(4.95) | 24.62(4.76) | 25.62(5.69) |
| Current smokers, % | 1119(12.2%) | 1788(12.7%) | 543(11.2%) | 689(12.2%) | 0 | 0 |
| CRP, mg/L^1^ | 1.40 (0.59,3.18) | 1.47(0.63,3.28) | 1.43(0.61,3.16) | 1.47(0.62,3.25) | 0.75(0.37,1.99) | 0.83(0.35,2.76) |
| IL6, pg/mL^2^ | 1.14(0.76,1.83) | 1.14(0.76,1.83) | 1.14(0.77,1.76) | 1.15(0.77,1.76) | 0.74(0.58,1.18) | 0.75(0.58,1.19) |

^1^ Sample size presented for the smallest and largest validations performed for the 1-way risk score (Table 5).

**Table S2: Number of metabolites available per NHS I, NHS II, HPFS sub-study.**

|  | **CRP risk score** | **IL6 risk score** |
| --- | --- | --- |
| **Metabolomics in risk score** | 319 | 232 |
| **Remove blank HMDB** | 234 | 176 |
| **NHS (n=12)** |  |  |
| nhs1.als | 51 | 38 |
| nhs1.breast | 52 | 38 |
| nhs1.colon | 50 | 39 |
| nhs1.diabetes | 58 | 45 |
| nhs1.exfoliation.glaucoma | 53 | 41 |
| nhs1.ibd | 63 | 46 |
| nhs1.ovarian | 63 | 47 |
| nhs1.parkinsons | 52 | 37 |
| nhs1.poag | 50 | 38 |
| nhs1.racial.diff | 52 | 38 |
| nhs1.rheumatoid | 64 | 46 |
| nhs1.stroke | 70 | 55 |
| **NHS II (n=8)** |  |  |
| nhs2.breast | 60 | 47 |
| nhs2.diabetes | 44 | 33 |
| nhs2.ibd | 62 | 46 |
| nhs2.ovarian | 62 | 48 |
| nhs2.poag | 51 | 38 |
| nhs2.rheumatoid | 64 | 47 |
| nhs2.stress | 62 | 47 |
| nhs2.stroke | 70 | 56 |
| **HPFS (n=5)** |  |  |
| hpfs.als | 54 | 36 |
| hpfs.colon | 51 | 37 |
| hpfs.exfoliation.glaucoma | 53 | 39 |
| hpfs.parkinsons | 56 | 37 |
| hpfs.poag | 52 | 36 |

**Table S3: Validation of the CRP and IL6 risk scores in NHS I, NHS II, and HPFS.** Minimal sample size – only individuals with both CRP and IL6 measurements.

|  |  | NHS I | |  | NHS II | |  | HPFS | |
| --- | --- | --- | --- | --- | --- | --- | --- | --- | --- |
|  | **R**  **(Pearson)** | ***P*** | **N** | **R**  **(Pearson)** | ***P*** | **N** | **R**  **(Pearson)** | ***P*** | **N** |
| CRP |  |  |  |  |  |  |  |  |  |
| ERS* | - | - | - | 0.478 | <0.001 | 311 | - | - | - |
| MRS** | 0.391 | <0.001 | 4035 | 0.427 | <0.001 | 564 | 0.208 | <0.001 | 537 |
| PRS | 0.446 | <0.001 | 8010 | 0.398 | <0.001 | 1702 | 0.429 | <0.001 | 2875 |
| MRS-ERS | - | - | - | 0.252 | 0.43 | 24 | - | - | - |
| PRS-MRS | 0.565 |  | 2292 | 0.510 | <0.001 | 174 | 0.437 | <0.001 | 376 |
| PRS-ERS | - | - | - | 0.539 | <0.001 | 170 | - | - | - |
| PRS-MRS-ERS | - | - | - | 0.346 | 0.33 | 12 | - | - | - |
| IL6 |  |  |  |  |  |  |  |  |  |
| ERS* | - | - | - | 0.327 | <0.001 | 121 | - | - | - |
| MRS** | 0.312 | <0.001 | 3163 | 0.354 | <0.001 | 673 | 0.199 | <0.001 | 605 |
| PRS | 0.698 | <0.001 | 5018 | 0.711 | <0.001 | 1401 | 0.690 | <0.001 | 1918 |
| MRS-ERS | - | - | - | 0.480 | 0.11 | 12 | - | - | - |
| PRS-MRS | 0.461 | <0.001 | 1886 | 0.521 | <0.001 | 181 | 0.397 | <0.001 | 464 |
| PRS-ERS | - | - | - | 0.437 | <0.001 | 86 | - | - | - |
| PRS-MRS-ERS | - | - | - | 0.668 | 0.035 | 12 | - | - | - |

*7 CpGs removed from the original 150-CpG ERS model due to differences in DNA methylation QC (removing probes)

**234 metabolites were removed from NHS, NHS II, and HPFS datasets due to the differences in the metabolomic platforms (blank HMDB).

**Table S20: Association between PRS, MRS, and PRS-MRS of inflammatory biomarkers and risk of all-cause mortality: CLSA**

|  | **Blood biomarker** | | **PRS** | | **MRS** | | **PRS-MRS** | |
| --- | --- | --- | --- | --- | --- | --- | --- | --- |
|  | **HR (95% CI)** | ***P*** | **HR (95% CI)** | ***P*** | **HR (95% CI)** | ***P*** | **HR (95% CI)** | ***P*** |
| **CRP** |  |  |  |  |  |  |  |  |
| Model 1 (age-adjusted) | 1.30 (1.24,1.37) | **<0.001** | 1.06 (1.01,1.12) | ***0.019*** | 1.33 (1.23,1.44) | **<0.001** | 1.32 (1.21,1.43) | **<0.001** |
| Model 2 (age, sex, smoking, race/ethnicity, alcohol) | 1.28 (1.19,1.37) | **<0.001** | 1.03 (0.96,1.10) | 0.42 | 1.44 (1.29,1.61) | **<0.001** | 1.37 (1.21,1.54) | **<0.001** |
| Model 3 (age, sex, smoking, race/ethnicity, alcohol, BMI) | 1.27 (1.18,1.36) | **<0.001** | 1.03 (0.96,1.10) | 0.44 | 1.44 (1.27, 1.64) | **<0.001** | 1.34 (1.17,1.53) | **<0.001** |
| **IL6** |  |  |  |  |  |  |  |  |
| Model 1 (age-adjusted) | 1.63 (1.47,1.80) | **<0.001** | 1.00 (0.92,1.01) | 0.911 | 1.57 (1.47,1.68) | **<0.001** | 1.39 (1.33,1.46) | **<0.001** |
| Model 2 (age, sex, smoking, race/ethnicity, alcohol) | 1.53 (1.34,1.75) | **<0.001** | 0.95 (0.85,1.07) | 0.44 | 1.61 (1.45, 1.79) | **<0.001** | 1.39 (1.31,1.48) | **<0.001** |
| Model 3 (age, sex, smoking, race/ethnicity, alcohol, BMI) | 1.50 (1.30,1.74) | **<0.001** | 0.95 (0.85, 1.07) | 0.43 | 1.65 (1.48,1.85) | **<0.001** | 1.39 (1.29,1.49) | **<0.001** |
| **Mutual adjustment + covariates from model 3:** | **Blood biomarker** | | **Risk score** | |  |  |  |  |
|  | **HR (95% CI)** | ***P*** | **HR (95% CI)** | ***P*** | **N** | **Events** |  |  |
| Blood CRP + CRP PRS + covariates | 1.30 (1.20,1.39) | **<0.001** | 0.94 (0.87,1.01) | 0.09 | 18105 | 790 |  |  |
| Blood IL6 + IL6 PRS + covariates | 1.49 (1.29,1.72) | **<0.001** | 0.94 (0.83,1.06) | 0.344 | 6591 | 265 |  |  |
| Blood CRP + CRP MRS + covariates | 1.06 (0.91,1.22) | 0.489 | 1.40 (1.20,1.64) | **<0.001** | 6855 | 290 |  |  |
| Blood IL6 + IL6 MRS + covariates | 1.22 (1.04,1.44) | ***0.015*** | 1.54 (1.34,1.77) | **<0.001** | 6728 | 274 |  |  |
| Blood CRP + CRP PRS-MRS + covariates | 1.09 (0.92,1.30) | 0.297 | 1.26 (1.05,1.51) | ***0.013*** | 6712 | 281 |  |  |
| Blood IL6 + IL6 PRS-MRS + covariates | 1.29 (1.11,1.51) | ***0.001*** | 1.33 (1.22,1.45) | **<0.001** | 6588 | 265 |  |  |

**Table S21: Association between PRS, MRS, and PRS-MRS of inflammatory biomarkers and risk of all-cause mortality: NHS+HPFS (combined)**

|  | **Blood biomarker** | | **PRS** | | **MRS** | | **PRS-MRS** | |
| --- | --- | --- | --- | --- | --- | --- | --- | --- |
|  | **HR (95% CI)** | ***P*** | **HR (95% CI)** | ***P*** | **HR (95% CI)** | ***P*** | **HR (95% CI)** | ***P*** |
| **CRP** |  |  |  |  |  |  |  |  |
| Model 1 (age-adjusted) | 1.19 (1.15, 1.22) | **<0.001** | 1.01 (0.98, 1.05) | 0.46 | 1.05 (1.01, 1.10) | ***2.90e-02*** | 1.02 (0.96, 1.08) | 0.59 |
| Model 2 (age, sex, smoking, race/ethnicity, alcohol) | 1.13 (1.07, 1.19) | **<0.001** | 0.98 (0.92, 1.03) | 0.48 | 1.06 (1.01, 1.12) | ***2.25e-02*** | 1.02 (0.96, 1.09) | 0.50 |
| Model 3 (age, sex, smoking, race/ethnicity, alcohol, BMI) | 1.08 (1.02, 1.15) | ***1.03e-02*** | 0.96 (0.90, 1.03) | 0.24 | 1.03 (0.98, 1.09) | 2.35e-01 | 0.99 (0.93, 1.06) | 0.79 |
| **IL6** |  |  |  |  |  |  |  |  |
| Model 1 (age-adjusted) | 1.27 (1.24, 1.31) | **<0.001** | 1.12 (1.08, 1.16) | **<0.001** | 1.11 (1.06, 1.16) | **<0.001** | 1.11 (1.05, 1.18) | ***<0.001*** |
| Model 2 (age, sex, smoking, race/ethnicity, alcohol) | 1.19 (1.13, 1.26) | **<0.001** | 1.09 (1.02, 1.16) | ***0.01*** | 1.10 (1.04, 1.15) | **<0.001** | 1.11 (1.04, 1.18) | ***1.47e-03*** |
| Model 3 (age, sex, smoking, race/ethnicity, alcohol, BMI) | 1.16 (1.10, 1.23) | **<0.001** | 1.05 (0.99, 1.13) | 0.13 | 1.06 (1.01, 1.12) | ***2.41e-02*** | 1.07 (1.00, 1.14) | 5.85e-02 |
| **Mutual adjustment + covariates from model 3:** | **Blood biomarker** |  | **Risk score** |  |  |  |  |  |
|  | **HR (95% CI)** | ***P*** | **HR (95% CI)** | ***P*** | **N** | **Events** |  |  |
| Blood CRP + CRP PRS + covariates | 1.15 (1.05, 1.25) | ***1.71e-03*** | 0.92 (0.85, 0.99) | **1.80e-02** | 7974 | 3657 |  |  |
| Blood IL6 + IL6 PRS + covariates | 1.20 (1.08, 1.33) | ***8.79e-04*** | 0.94 (0.85, 1.03) | 0.20 | 7974 | 3657 |  |  |
| Blood CRP + CRP MRS + covariates | 1.08 (1.01, 1.14) | ***2.25e-02*** | 1.01 (0.96, 1.07) | 0.66 | 3905 | 1966 |  |  |
| Blood IL6 + IL6 MRS + covariates | 1.15 (1.09, 1.21) | ***8.97e-07*** | 1.03 (0.98, 1.09) | 0.21 | 3905 | 1966 |  |  |
| Blood CRP + CRP PRS-MRS + covariates | 1.14 (1.05, 1.25) | ***3.02e-03*** | 0.94 (0.87, 1.01) | 9.28e-02 | 2278 | 1219 |  |  |
| Blood IL6 + IL6 PRS-MRS + covariates | 1.12 (1.04, 1.21) | ***2.95e-03*** | 1.03 (0.96, 1.10) | 0.49 | 2278 | 1219 |  |  |

**Table S22: Association between PRS, MRS, and PRS-MRS of inflammatory biomarkers and risk of all-cause mortality, restricted to 65y and above: CLSA**

|  | **Blood biomarker** | | **PRS** | | **MRS** | | **PRS-MRS** | |
| --- | --- | --- | --- | --- | --- | --- | --- | --- |
|  | **HR (95% CI)** | ***P*** | **HR (95% CI)** | ***P*** | **HR (95% CI)** | ***P*** | **HR (95% CI)** | ***P*** |
| **CRP** |  |  |  |  |  |  |  |  |
| Model 1 (age-adjusted) | 1.25 (1.18,1.32) | **<0.001** | 1.06 (1.00,1.13) | ***0.027*** | 1.26 (1.16,1.38) | **<0.001** | 1.27 (1.16,1.40) | **<0.001** |
| Model 2 (age, sex, smoking, race/ethnicity, alcohol) | 1.26 (1.09,1.13) | **<0.001** | 1.05 (0.97,1.13) | 0.24 | 1.36 (1.19,1.56) | **<0.001** | 1.32 (1.15,1.52) | **<0.001** |
| Model 3 (age, sex, smoking, race/ethnicity, alcohol, BMI) | 1.21 (1.12,1.32) | **<0.001** | 1.03 (0.95,1.11) | 0.50 | 1.30 (1.11,1.52) | **<0.001** | 1.26 (1.08,1.46) | ***0.002*** |
| **IL6** |  |  |  |  |  |  |  |  |
| Model 1 (age-adjusted) | 1.51 (1.36,1.68) | **<0.001** | 1.00 (0.91,1.11) | 0.99 | 1.50 (1.38,1.63) | **<0.001** | 1.50 (1.38,1.63) | **<0.001** |
| Model 2 (age, sex, smoking, race/ethnicity, alcohol) | 1.40 (1.21,1.63) | **<0.001** | 0.94 (0.82,1.08) | 0.37 | 1.53 (1.33,1.76) | **<0.001** | 1.53 (1.33,1.76) | **<0.001** |
| Model 3 (age, sex, smoking, race/ethnicity, alcohol, BMI) | 1.35 (1.15,1.58) | **<0.001** | 0.94 (0.82,1.08) | 0.39 | 1.51 (1.29,1.78) | **<0.001** | 1.50 (1.28,1.76) | **<0.001** |
| **Mutual adjustment + covariates from model 3:** | **Blood biomarker** | | **Risk score** | |  |  |  |  |
|  | **HR (95% CI)** | ***P*** | **HR (95% CI)** | ***P*** | **N** | **Events** |  |  |
| Blood CRP + CRP PRS + covariates | 1.23 (1.13,1.34) | **<0.001** | 0.96 (0.88,1.04) | 0.34 | 6854 | 616 |  |  |
| Blood IL6 + IL6 PRS + covariates | 1.32 (1.13,1.55) | **<0.001** | 0.93 (0.81,1.07) | 0.33 | 2479 | 197 |  |  |
| Blood CRP + CRP MRS + covariates | 1.04 (0.87,1.23) | 0.69 | 1.27 (1.04,1.54) | **0.016** | 2600 | 219 |  |  |
| Blood IL6 + IL6 MRS + covariates | 1.14 (0.95,1.37) | 0.15 | 1.41 (1.16,1.70) | **<0.001** | 2529 | 205 |  |  |
| Blood CRP + CRP PRS-MRS + covariates | 1.03 (0.85,1.25) | 0.76 | 1.23 (1.00,1.51) | **0.047** | 2548 | 211 |  |  |
| Blood IL6 + IL6 PRS-MRS + covariates | 1.11 (0.92,1.34) | 0.27 | 1.43 (1.18,1.72) | **<0.001** | 2479 | 197 |  |  |

**Table S23: Association between PRS, MRS, and PRS-MRS of inflammatory biomarkers and risk of all-cause mortality, restricted to 65y and above: NHS+HPFS (combined)**.

|  | **Blood biomarker** | | **PRS** | | **MRS** | | **PRS-MRS** | |
| --- | --- | --- | --- | --- | --- | --- | --- | --- |
|  | **HR** | ***P*** | **HR** | ***P*** | **HR** | ***P*** | **HR** | ***P*** |
| **CRP** |  |  |  |  |  |  |  |  |
| Model 1 (age-adjusted) | 1.13 (1.08, 1.19) | **<0.001** | 1.01 (0.95, 1.07) | 0.81 | 1.04 (0.95, 1.14) | 0.41 | 1.02 (0.91, 1.13) | 0.76 |
| Model 2 (age, sex, smoking, race/ethnicity, alcohol) | 1.06 (0.96, 1.16) | 0.27 | 0.96 (0.85, 1.08) | 0.46 | 1.02 (0.92, 1.12) | 0.72 | 1.02 (0.90, 1.14) | 0.80 |
| Model 3 (age, sex, smoking, race/ethnicity, alcohol, BMI) | 1.04 (0.94, 1.16) | 0.45 | 0.95 (0.84, 1.08) | 0.45 | 1.00 (0.91, 1.11) | 0.99 | 1.00 (0.88, 1.13) | 0.97 |
| **IL6** |  |  |  |  |  |  |  |  |
| Model 1 (age-adjusted) | 1.22 (1.16, 1.28) | **<0.001** | 1.12 (1.06, 1.19) | **<0.001** | 1.16 (1.06, 1.27) | **1.31e-03** | 1.19 (1.07, 1.32) | **1.53e-03** |
| Model 2 (age, sex, smoking, race/ethnicity, alcohol) | 1.19 (1.07, 1.31) | **0.001** | 1.16 (1.03, 1.29) | **1.26e-02** | 1.12 (1.02, 1.23) | **2.14e-02** | 1.17 (1.05, 1.30) | **5.89e-03** |
| Model 3 (age, sex, smoking, race/ethnicity, alcohol, BMI) | 1.20 (1.08, 1.33) | **0.001** | 1.13 (1.00, 1.27) | **4.45e-02** | 1.10 (1.00, 1.21) | 6.13e-02 | 1.13 (1.01, 1.27) | **3.08e-02** |
| **Mutual adjustment + covariates from model 3:** | **Blood biomarker** | | **Risk score** | |  |  |  |  |
|  | **HR** | ***P*** | **HR** | ***P*** | **N** | **Events** |  |  |
| Blood CRP + CRP PRS + covariates | 1.06 (0.92, 1.22) | 0.42 | 0.93 (0.81, 1.07) | 0.30 | 1431 | 758 |  |  |
| Blood IL6 + IL6 PRS + covariates | 1.19 (0.98, 1.44) | 8.07e-02 | 1.01 (0.85, 1.20) | 0.89 | 1431 | 758 |  |  |
| Blood CRP + CRP MRS + covariates | 1.04 (0.94, 1.16) | 0.46 | 0.99 (0.89, 1.10) | 0.87 | 629 | 353 |  |  |
| Blood IL6 + IL6 MRS + covariates | 1.18 (1.07, 1.30) | **1.21e-03** | 1.07 (0.98, 1.18) | 0.15 | 629 | 353 |  |  |
| Blood CRP + CRP PRS-MRS + covariates | 1.04 (0.90, 1.20) | 0.62 | 0.98 (0.85, 1.13) | 0.77 | 424 | 236 |  |  |
| Blood IL6 + IL6 PRS-MRS + covariates | 1.14 (1.00, 1.30) | **4.30e-02** | 1.09 (0.97, 1.23) | 0.16 | 424 | 236 |  |  |

**List of additional Supplemental Tables in a separate Excel file:**

**Table S3: 1-way ERS prediction models**

**Table S4: 1-way MRS prediction models**

**Table S5: 1-way PRS prediction models**

**Table S6: Two-way models**

**Table S7: Three-way models**

**Table S8: Minimal sample size validation of CRP and IL6 scores**

**Table S9: All mortality models CRP**

**Table S10: All mortality models IL6**

**Table S11: All mortality models TNFa**

**Table S12: Sensitivity analyses: 1-way ERS**

**Table S13: Sensitivity analyses: 1-way MRS**

**Table S14: Sensitivity analyses: 1-way PRS**

**Table S15: Sensitivity analyses: 2-way PRS-MRS**

**Table S16: Sensitivity analyses: 2-way PRS-ERS**

**Table S17: Sensitivity analyses: 2-way MRS-ERS**

**Table S18: Sensitivity analyses: 3-way PRS-MRS-ERS**

**Table S19: Multiple mutual adjustments**

**Figure S1: Correlation of age with inflammation markers in the CLSA study.** (a) CRP; (b) IL6; (c) TNFa.

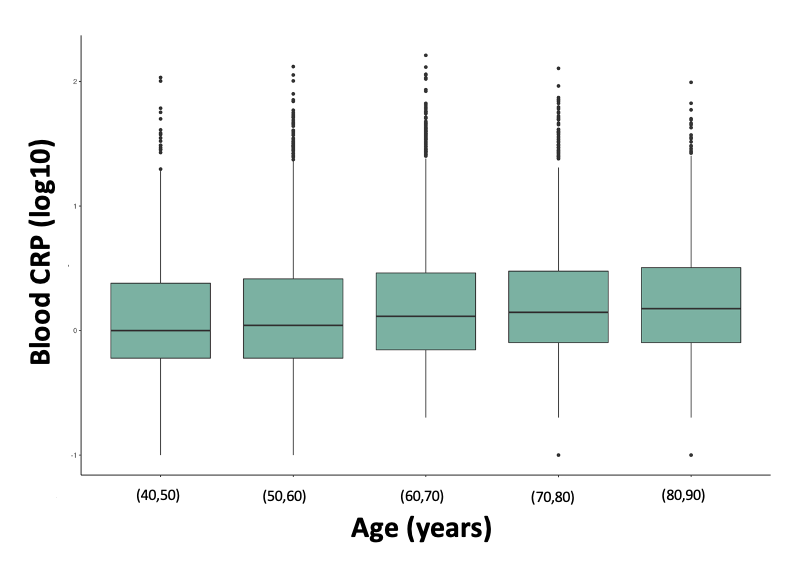

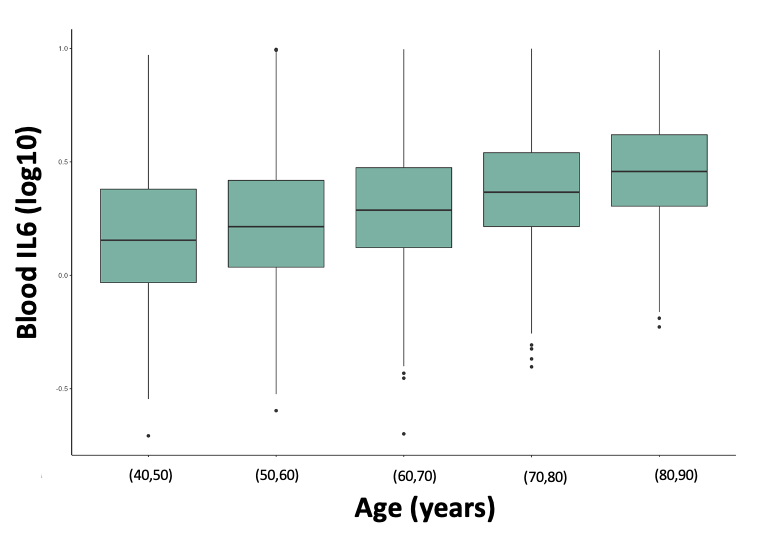

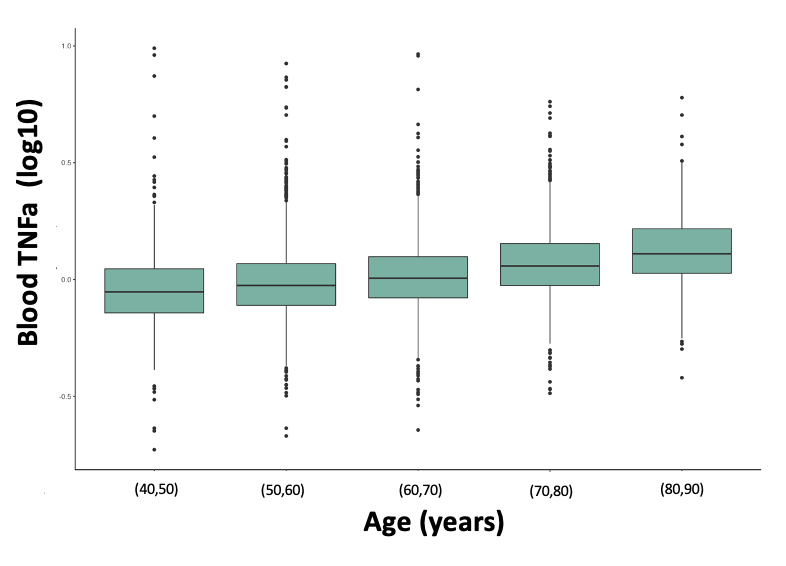

**Figure S2: 1-way risk scores of inflammation markers: Genetics via PRS, the CLSA study.** (a) CRP; (b) IL6; (c) TNFa.

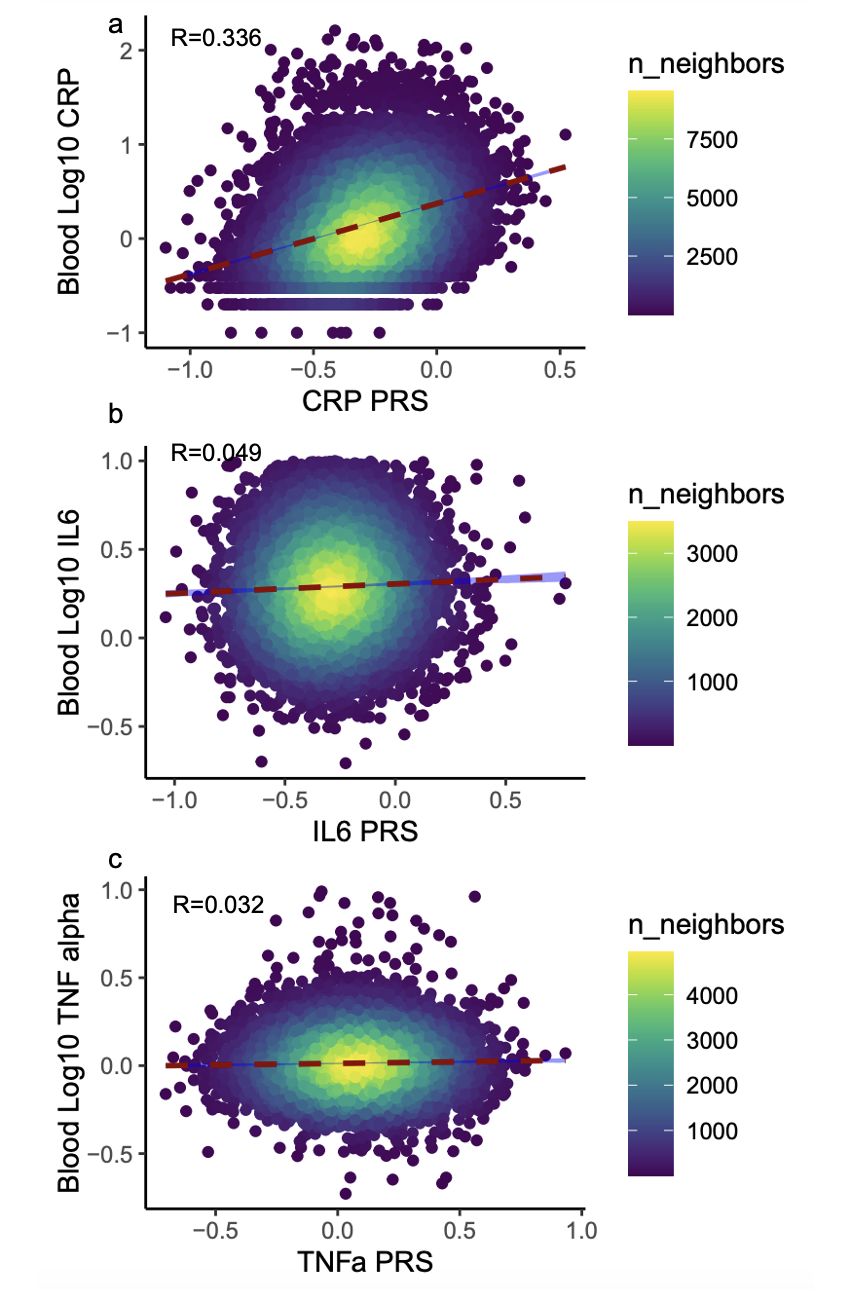

**Figure S3:** **1-way risk scores of inflammation markers: MRS.** (a) CRP; (b) IL6; (c) TNFa. Data was batch-effect normalized and standardized; metabolomics outliers were removed. Undefined metabolites were removed. The metabolites at each fold ranged between 188-400 for CRP, 165-305 for IL6, and 95-261 for TNFa. Metabolite distribution used for the prediction model applied to the validation sets is also presented below.

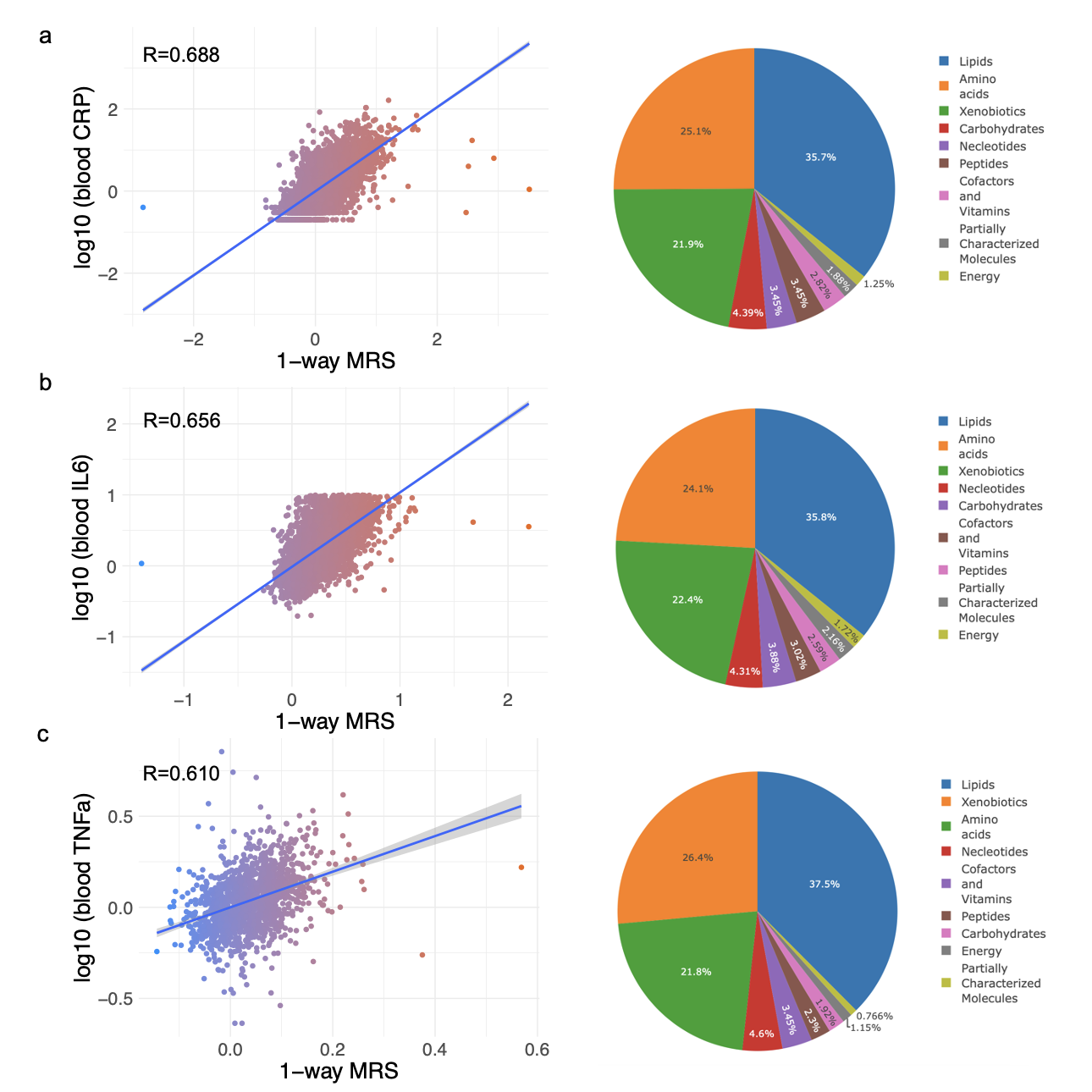

**Figure S4: 1-way risk score of inflammation markers: ERS (DNA methylation).** (a) CRP; (b) IL6; (c) TNFa. The CpGs at each fold ranged between 122-328 for CRP, 356-527 for IL6, and 137-769 for TNFa.

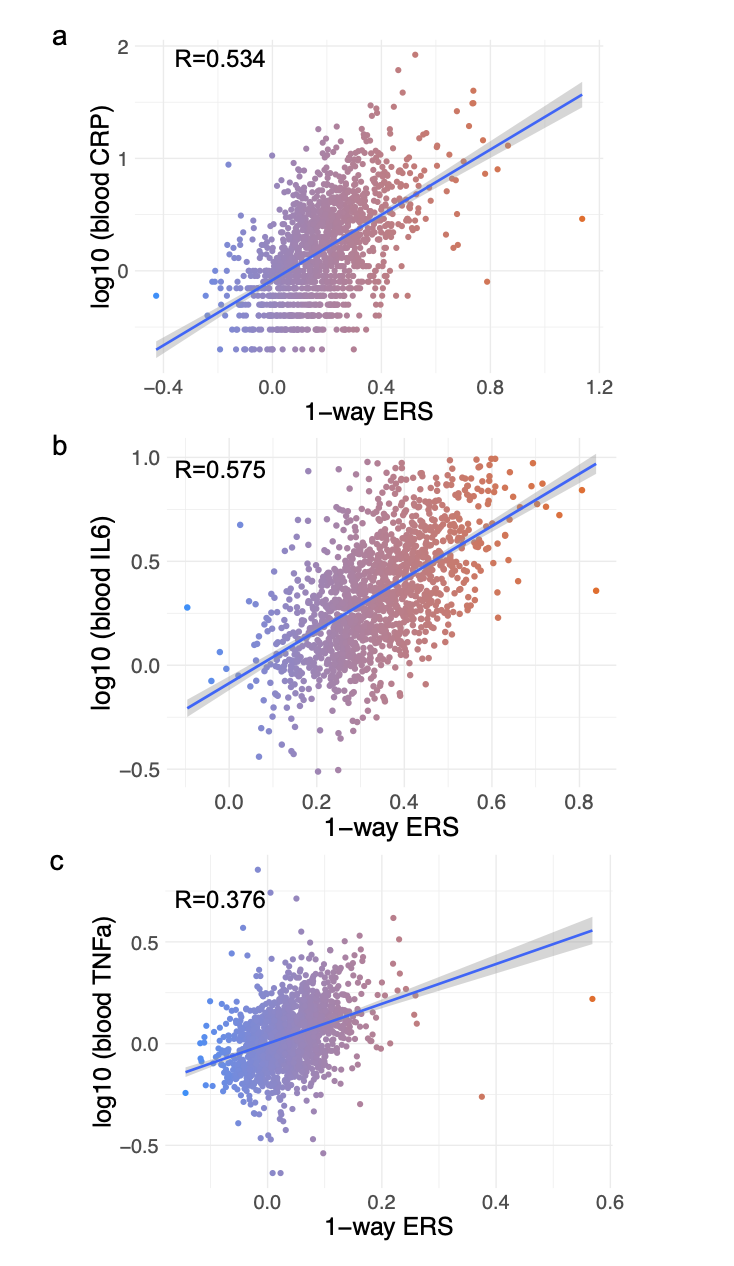

**Figure S5: Dynamic Area Under the Curve (AUC) of the survival models**. a. 3-way CRP risk score: inputting traditional markers first and 3-way risk score last (AUC=0.744); b. 3-way IL6 risk score: inputting traditional markers first and 3-way risk score last (AUC=0.772); c. 3-way TNFa risk score: inputting traditional markers first and 3-way risk score last (AUC=0.789).

a.
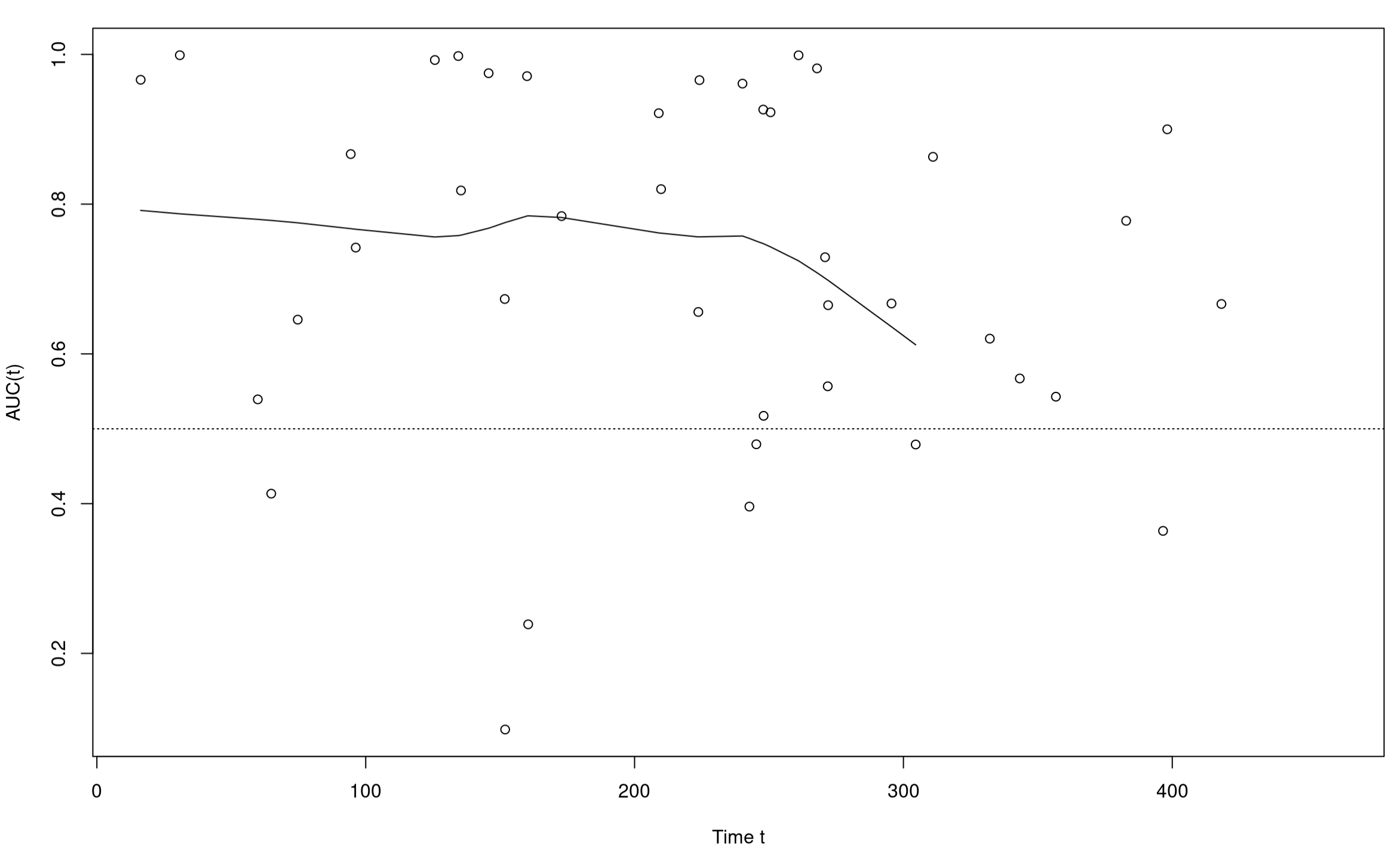
 b.
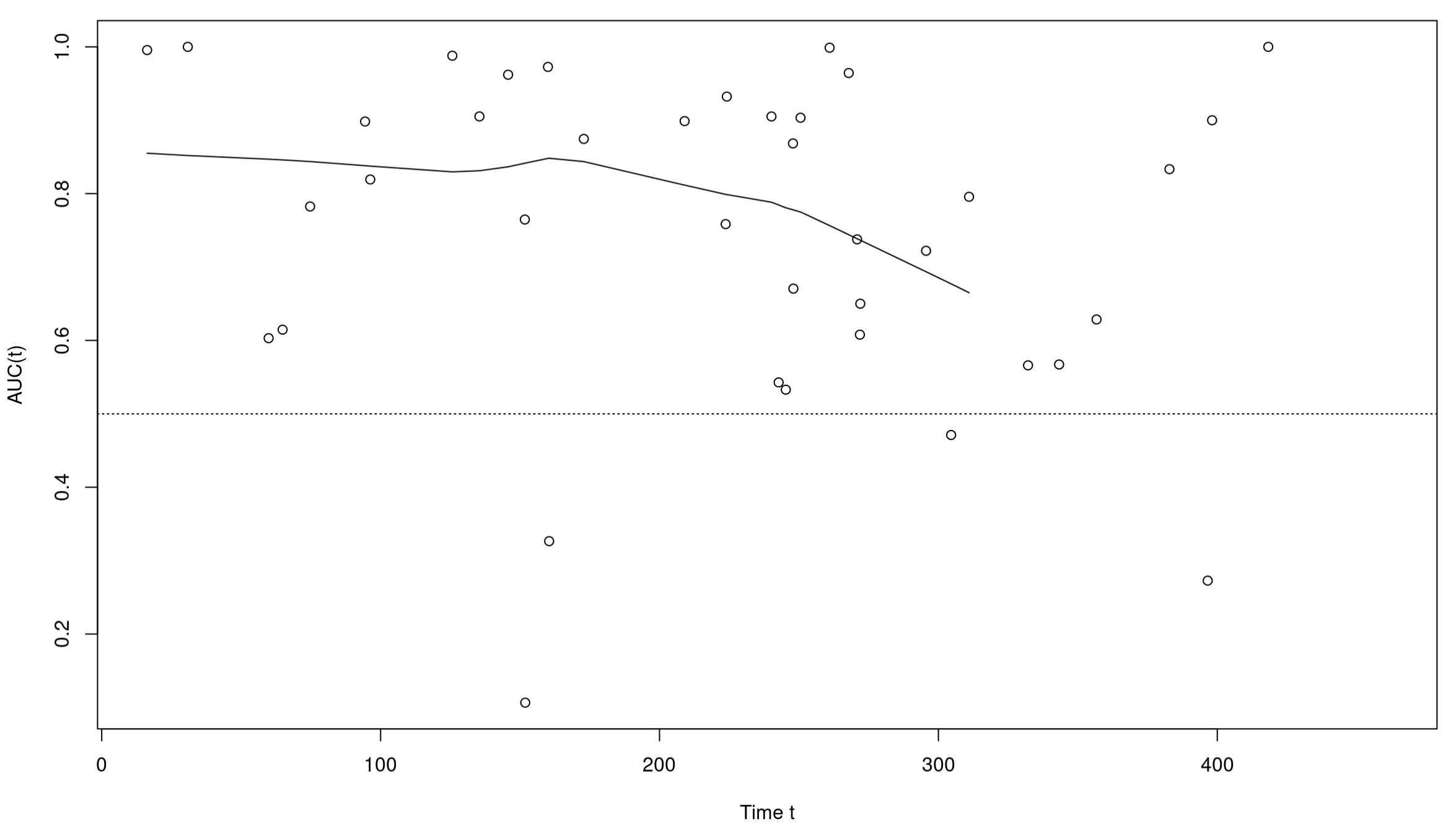

c.
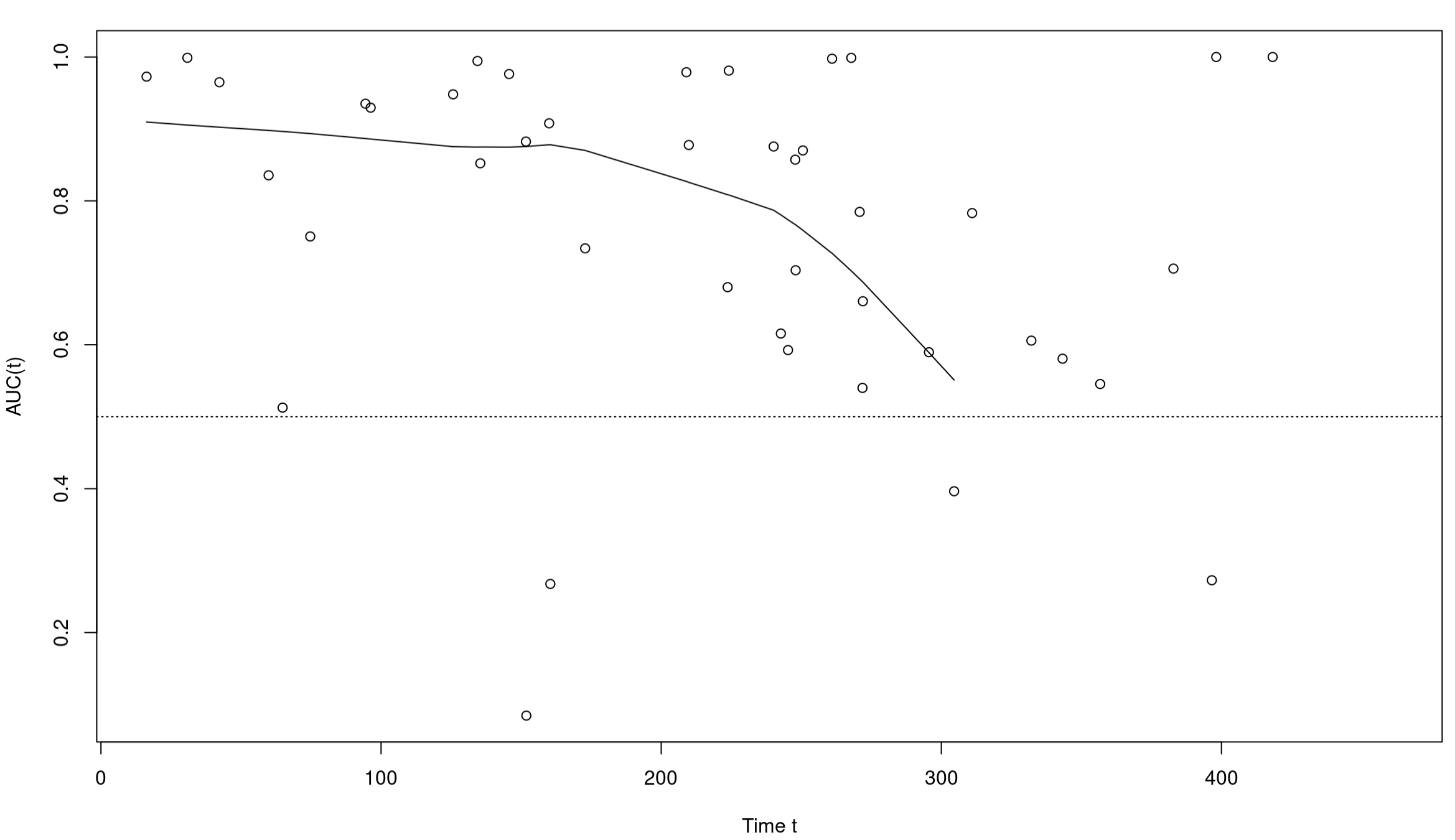

1.	Colditz, G. A., Manson, J. E. & Hankinson, S. E. The Nurses’ Health Study: 20-year contribution to the understanding of health among women. *J Womens Health* 6, 49–62 (1997).

2.	Rimm, E. B. *et al.* Prospective study of alcohol consumption and risk of coronary disease in men. *The Lancet* 338, 464–468 (1991).

3.	Li, J. *et al.* The Mediterranean diet, plasma metabolome, and cardiovascular disease risk. *Eur Heart J* 41, 2645 (2020).

4.	Li, J. *et al.* Dietary inflammatory potential and risk of cardiovascular disease among men and women in the US. *J Am Coll Cardiol* 76, 2181–2193 (2020).
